# A model to analyze rideshare data to surveil novel strains of SARS-CoV-2

**DOI:** 10.1101/2021.05.07.21256856

**Authors:** Conrad W. Safranek, David Scheinker

## Abstract

**Background:** The emergence of novel, potentially vaccine-resistant strains of SARS-CoV-2 poses a serious risk to public health. The interactions between passengers and drivers facilitated by rideshare platforms such as Uber are, essentially, a series of partially standardized, random experiments of SARS-CoV-2 transmission. Rideshare companies share data with government health agencies, but no statistical method is available to aggregate these data for the systematic study of the transmission dynamics of COVID-19.

**Methods:** We develop a proof-of-concept model for the analysis of data from rideshare interactions merged with COVID-19 diagnosis records. Using simulated data with rideshare volumes, disease prevalence, and diagnosis rates based on a large US city, we use the model to test hypotheses about the emergence of viral strains and their transmission characteristics in the presence of non-pharmaceutical interventions and superspreaders.

**Findings:** Data from 10 simulated trials of SARS-CoV-2 propagation within the Los Angeles rideshare network resulted in an average of 190,387.1 potentially infectious rideshare interactions. Assuming access to data on 25% of the total estimated infections (*Partial Reporting*), these interactions resulted in an average of 409.0 diagnosed rideshare infections given our transmission model assumptions. For each of the 10 simulated trials, analysis given *Partial Reporting* could consistently differentiate between a baseline strain and an emergent, more infectious viral strain, enabling hypothesis testing about transmission characteristics.

**Interpretation:** Simulated evaluation of a novel statistical model suggests that rideshare data combined with COVID-19 diagnosis data have the potential to automate continued surveillance of emergent novel strains of SARS-CoV-2 and their transmission characteristics.

## INTRODUCTION

The emergence of novel, potentially vaccine-resistant strains of SARS-CoV-2 poses a serious threat to ongoing efforts to public health.^1^ Policies to contain the spread of COVID-19 rely on understanding what such strains mean for the risk of transmission and how they may impact optimal strategies to contact trace, test, quarantine, and vaccinate.^2–6^ Important factors include the infectivity of new viral strains, the onset of infectivity relative to symptom onset, the use and effectiveness of non-pharmaceutical interventions (NPIs) such as facemasks and hand sanitizer, and the prevalence of superspreaders in the population.^5,7,8^ Beyond SARS-CoV-2, such considerations will remain relevant for future pandemics.

The study of the transmission of SARS-CoV-2 has been constrained primarily to specialized settings such as laboratory experimentation, by examining infections in ships, hospitals, and military facilities, and through placebo-controlled vaccine clinical trials.^9–14^ The usefulness of these data is limited by the difficulty of translating the results to other settings and the emergence of new viral strains. Practical and ethical limitations to clinical trials and the challenges of drawing causal inference from observational data limit the understanding of the real-world transmission dynamics of COVID-19. Mobile phone applications for contact tracing gather real-world data, but with limited information about the setting in which the interaction took place. Aerosolized viral dynamics within cars have been studied,^15^ and rideshare companies already share data with government health agencies for contact tracing,^16^ but to the best of our knowledge no statistical method is available for the use of these data for the systematic study of the transmission dynamics of COVID-19.

The interactions between passengers and drivers facilitated by rideshare platforms such as Uber and Lyft are, essentially, a series of partially controlled, standardized, random experiments of SARS-CoV-2 transmission. Rideshare trips are often the only connection between individuals; are governed by mask-wearing policies implemented on specific dates; occur in a relatively controlled environment with fairly consistent spatial dynamics; and are sporadic with respect to time, location, and duration. Rideshare location and time data are stored in a machine-readable format that could be linked to data such as diagnoses or self-reported symptoms (anonymized in accordance with local legislation). Rideshare trip data may facilitate the automated identification of the emergence of new viral strains and the study of their transmission dynamics, with and without the presence of NPIs, and based on passenger and driver characteristics.

The unknown and potentially low “signal-to-noise” ratio of detected-to-undetected cases in a rideshare network presents a challenge for the usefulness of a statistical method of rideshare data merged with diagnoses data.^17^ The potential utility of such a method depends on its performance detecting rideshare-acquired infections while accounting for “false positives” rideshare interactions that appear to have resulted in transmission even though the potential infectee contracted the virus elsewhere.

We develop Rideshare Infection Detection (RIDE), a probabilistic model of rideshare transmissions designed to test hypotheses about the emergence of novel strains of SARS-CoV-2 and their transmission dynamics. Since such aggregated data are not available, we simulate viral transmission data in a hypothetical rideshare network based on empirical data from a large US city. We use these simulated data to test hypotheses about transmission dynamics, while assuming access to only the kinds of data that may be available in practice.

## METHODS

### Overview

We use an established mathematical model of viral transmission, adapted to rideshare interactions, to estimate the probability of a rideshare trip resulting in infection. We use empirical data from Los Angeles (LA) COVID-19 records and rideshare-use statistics during quarantine to generate synthetic rideshare infection patterns by simulating the propagation of the virus through a rideshare network. We use these synthetic data to evaluate the power of RIDE to test hypotheses regarding the factors most influential for COVID-19 disease spread.

### Mathematical model of transmission

We estimate patient infectivity relative to symptom onset with a general mathematical model of viral transmission adapted with SARS-CoV-2-specific parameter estimates from the literature.^9,18,19^ Probability-of-infection functions incorporating ride duration and ride timing relative the infector’s symptom onset are derived for unique passenger-to-driver and driver-to-passenger transmission dynamics. We define four hypothetical scenarios of transmission (Table 1). Scenario *viral variant A* assumes facemasks are not used and that there are no superspreaders. This serves as the baseline. In scenario *viral variant A with masks*, we represent masking with a 50% reduction in viral particle exchange between infector and potential infectee. In scenario *viral variant A with superspreaders*, we introduce infectivity asymmetry in the population, with one in 20 infected individuals having a 5-fold higher probability of transmitting SARS-CoV-2.^20^ In scenario *viral variant B*, based on research detailing the emergent D614G SARS-CoV-2 strain, we introduce a viral variant with 300% higher viral load relative to the baseline *viral variant A* (Supplementary section II).^7^

**Table 1.** Hypothesized sets of SARS-CoV-2 transmission parameters and probability of infection for populations with differing transmission characteristics.

| Parameters | Estimates from literature<br>– mean [95% confidence interval] | Viral variant A |  |  | Viral variant B |
| --- | --- | --- | --- | --- | --- |
|  |  | No masks, no super-spreaders | Masks, no super-spreaders† | No masks, 5% super-spreaders‡ | No masks, no super-spreaders |
| **Viral expulsion** relative to viral variant A – % | *Not available* | 100% | - | - | 300% |
| Viral exchange reduction due to **NPIs** – % | *Not available* | - | 50% | - | - |
| Infectivity of **superspreader** relative to baseline viral variant A – % | *Not available* | - | - | 500% | - |
| Days infectious before symptom onset – days | 2.3 [0.8-3.0] | 2.3 | 2.3 | 2.3 | 3 |
| Peak infectivity relative to symptom onset – days * | -0.7 [-2.0-0.2] | -0.7 | -0.7 | -0.7 | -2 |
| probability of **passenger-to-driver** infection from 20 min ride w/ passenger on day of symptom onset – % | - | 1.5% | 0.7% | 7.40% | 2.8% |
| probability of **driver-to-passenger** infection from 20 min ride w/ driver on day of symptom onset – % | - | 2.9% | 1.5% | 14.70% | 5.6% |

\*Adjusted for via gamma infectivity distribution shape and scale parameters. †Passenger and driver both wearing masks, blocking 50% of viral particle exchange. ‡Assumes 5% of infected individuals are superspreaders and 95% of infected individuals are non-superspreaders; non-superspreaders are 78.9% as infectious as baseline individuals with *viral variant A*.

### Synthetic rideshare transmission data

We generate synthetic data to represent the data that may become available if public health agencies partner with rideshare companies. The characteristics of urban rideshare networks, COVID-19 diagnoses, and estimates of the fraction of cases represented by the diagnoses from March 17th to October 3rd (200 days) in Los Angeles county (LA) were derived from the literature (Supplementary section III).^21–23^ LA rideshare volume, with an estimated 75% reduction during periods of quarantine, was simulated.^23^ The total number of infections in LA was assumed to be twice the number of infections reported over the period considered.^24,25^ The number of infections as a fraction of the population of LA was used as the probability that each person in the simulation was infected outside the rideshare network. Each network was initialized by assigning a day of symptom onset to randomly selected passengers and drivers, with the number of infections proportional to the estimated historical number of infections reported 3 days later, to account for the average delay between symptom onset and testing.^26^ The time of symptom onset was chosen from a normal distribution calibrated based on empirical LA diagnoses data.

For each interaction involving an infected individual, the probability of infection, passenger-to-driver or driver-to-passenger, was calculated using the parameter set corresponding to the viral scenario being tested. The interaction results in an infection based on a draw from a Bernoulli random variable with probability of success equal to the calculated probability of infection. For those infected, an incubation period was drawn from a probability distribution and subsequent symptom onset time assigned (Supplementary section III).

From simulation, we derive data representative of the empirical data that would be available to an epidemiologist. We consider a *Partial Reporting* scenario with access to diagnosis and symptom onset data for 25% of infections and a *Full Reporting* scenario in which data are available for all infections (Supplementary section III).

### Hypothesis testing with RIDE

Using only the data that would be available to an epidemiologist (neither undiagnosed infections nor information on the origin of each diagnosed infection), we introduce Rideshare Infection Detection (RIDE) to analyze the simulated rideshare infection patterns in order to calculate the number of expected infections and the number of observed infections (Figure 1).

**Figure 1.**
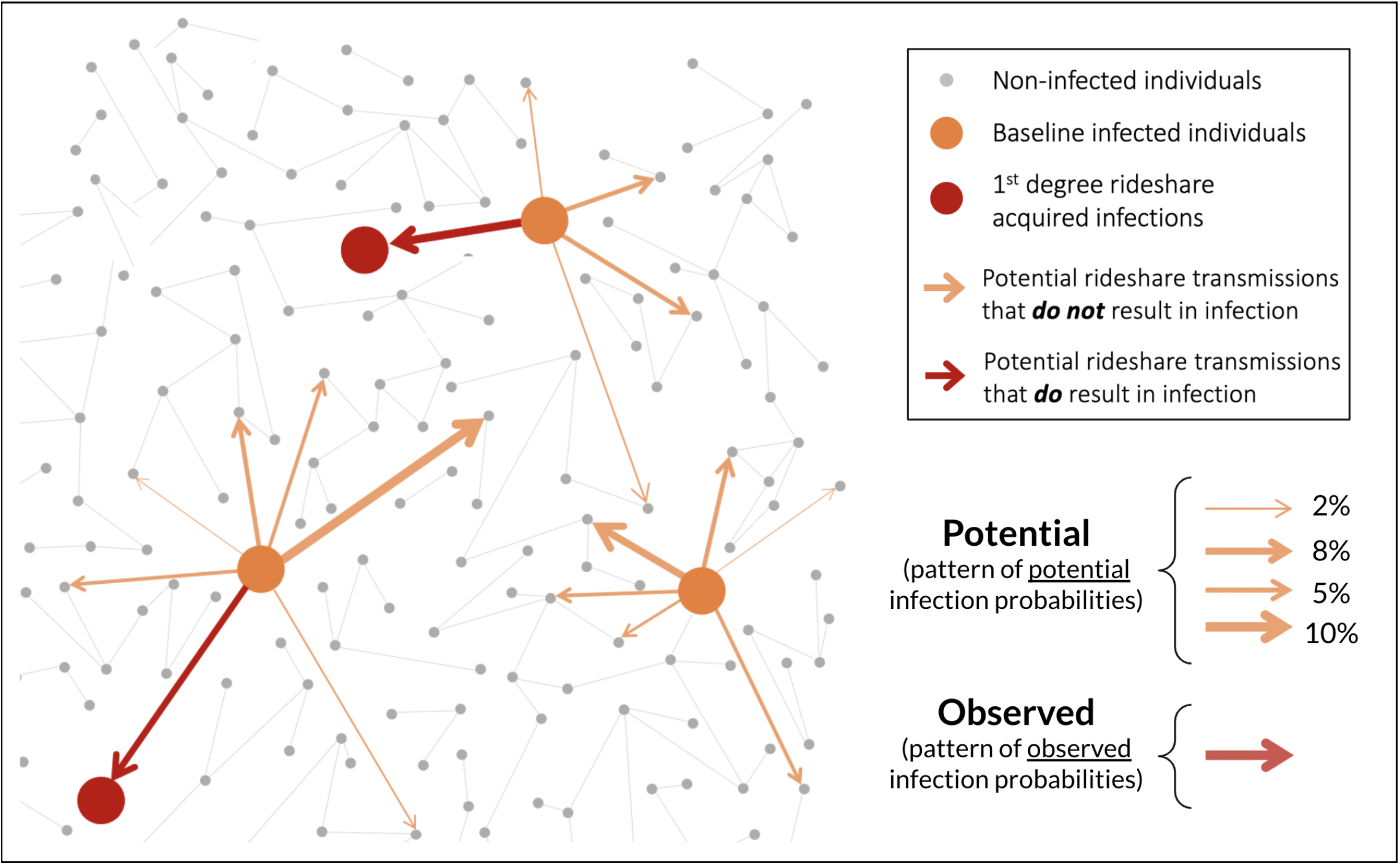
Simplified schematic of RIDE analytical method to identify potential and observed infections. Observed infections are tabulated by counting individuals with symptom onset following a potentially infectious rideshare interaction. The expected number of infections is the sum of all potential infections, each weighted by its probability. Arrow thickness for each potential infection corresponds to the probability-of-infection magnitude, which is calculated given a mathematical model of rideshare transmission that depends on ride duration, the timing of the ride relative to the potential infector’s symptom onset, and the assumed SARS-CoV-2 transmission parameters given the hypothesized scenario and viral variant.

We use a simulation to generate data using the parameters for *viral variant A* and analyze the results under the assumption that the data were generated with parameters corresponding to each scenario considered.

Using the synthetic data produced via simulated propagation of *viral variant A*, we test hypotheses about which scenario (*viral variant A, viral variant A with masks*, or *viral variant B*) corresponds to the number of apparent rideshare infections in the network. For each scenario, we use the parameter set corresponding to the scenario being tested and knowledge only of diagnosed infections to calculate the *Expected* number of rideshare infections, equal to the sum of the probabilities of infection across all potentially infectious rideshare interactions (i.e. the sum of the expected values of the Bernoulli distributions). The observed number of infections is determined by counting the number of interactions in which a diagnosed individual within their infectious window shared a rideshare vehicle with a potential infectee who had a positive diagnosis with symptom onset between 1.5 and 10 days following the rideshare trip. This observed number is adjusted to account for the percentage of cases diagnosed (this diagnosis percentage is assumed known) and for an estimation of the average number of “false positives”—interactions that appeared to have resulted in an infection even though the infectee was infected elsewhere—given the overall infection density in the network (Supplementary section IV). For each respective level of reporting (*Full* vs. *Partial Reporting)*, this simulated propagation and analysis with RIDE was repeated 10 times to determine the impact of transmission stochasticity and rideshare network variability. Across the 10 simulations, the differences in the expected number of passenger-to-driver infections (given each set of hypothesized parameters) and the observed number of passenger-to-driver infections are compared with a pairwise Kruskal-Wallis test.

Separately, two synthetic rideshare infection patterns were simulated and compared: propagation of *viral variant A* and propagation of *viral variant A with superspreaders*. For each round of analysis, the difference was calculated between the number of observed passengers infected per infectious driver in the superspreader scenario minus the non-superspreader scenario. This was repeated 10 times each for *Partial* and *Full Reporting* and the resulting distributions were compared with the Kruskal-Wallis test. All p-values were adjusted for multiple testing.

All simulated viral propagation and analysis with RIDE were performed with R (Version 4.0.3, 2010; Vienna, Austria), and executed with Stanford’s Sherlock High-Performance Computing Cluster.

## RESULTS

### Mathematical model

Differences in viral strains and the alternative hypothetical propagation scenarios lead to significant differences in the probability of rideshare SARS-CoV-2 infection. Relative to the baseline scenario (*viral variant A*) probability of infection given a 20 minute ride with a passenger one day before symptom onset, the probability of the driver being infected is 51% lower when both driver and passenger are masked (scenario *viral variant A with masks*), 480% higher if the passenger is one of the more infectious individuals from scenario *viral variant A with superspreaders*, and 342% higher if the passenger has *viral variant B* (Figure 2).

**Figure 2.**
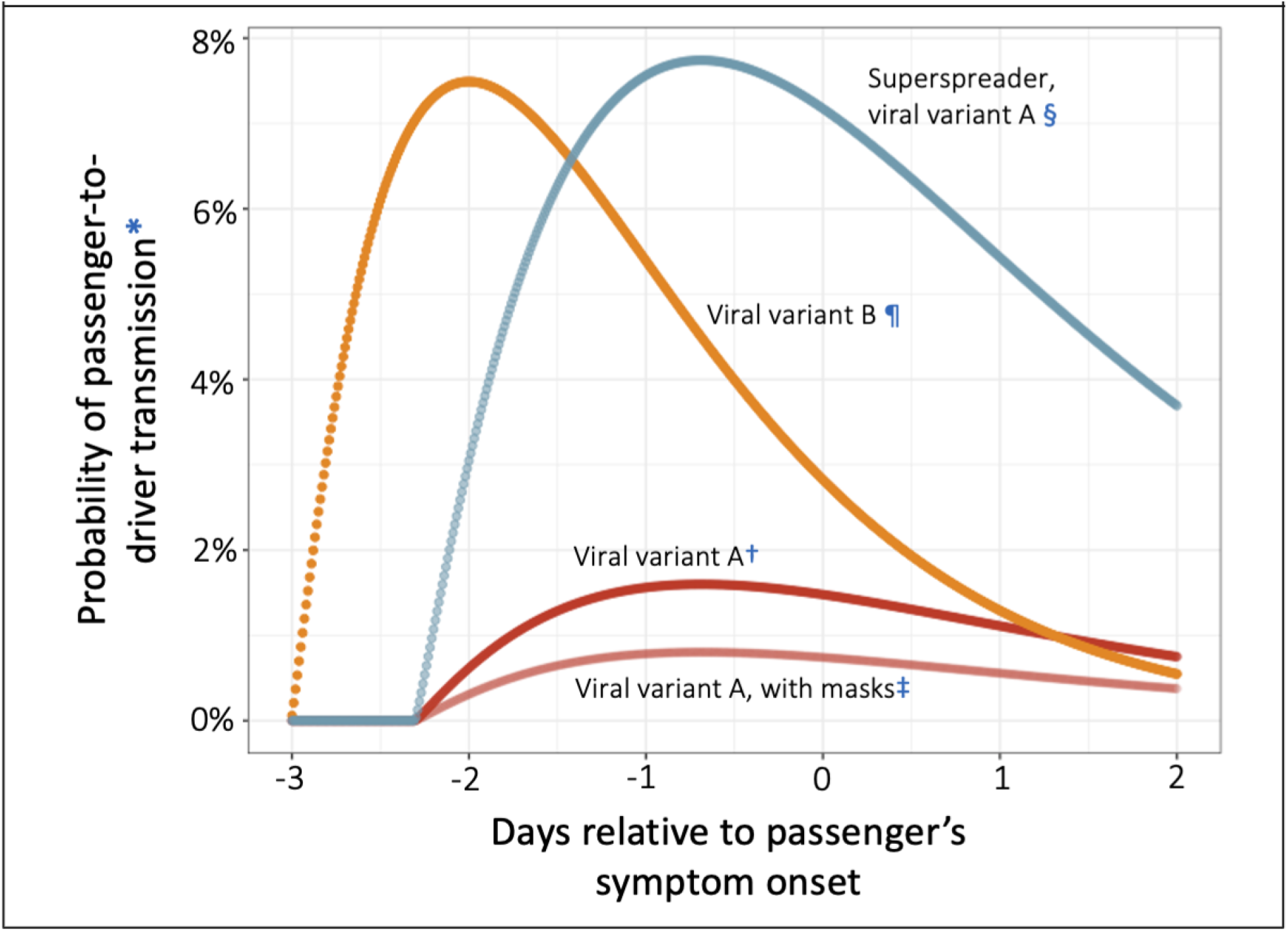
Modeled probability of transmission from passenger to driver. Probability of infection changes significantly depending upon interaction dynamics (time of ride relative to passenger’s symptom onset, and ride duration) and upon assumptions defining the hypothetical SARS-CoV-2 transmission parameters (representing the passenger’s viral variant, whether the passenger is a superspreader, and whether face masks are used). *Probability of driver infection given 20 min ride with infected passenger. †Baseline *viral variant*. ‡Passenger and driver wearing masks, blocking 50% of viral particle exchange. §Superspreaders with *viral variant* A are 500% more infectious than baseline individuals with viral variant A. ¶Alternative *viral variant B* is 300% more infectious than baseline and has different infectivity parameters within the previously estimated 95% confidence intervals.

### Synthetic rideshare transmission data

The simulation was initiated with a baseline infection probability of 528,828 out of 10 million, twice the 264,414 reported infections in LA county during this period. Data from 10 simulated trials of SARS-CoV-2 propagation within the Los Angeles rideshare network resulted in an average of 190,387.1 (range 187,898 to 193,645) potentially infectious rideshare interactions, encompassing possible passenger-to-driver and driver-to-passenger transmissions. When these ten simulated trials were propagated assuming SARS-CoV-2 transmission parameters corresponding to *viral variant A*, there were an average of 409.0 (range 384 to 424) rideshare infections resulting in a diagnosis for the *Partial Reporting* scenario, and 1,666.4 (range 1614 to 1698) rideshare infections for the *Full Reporting* scenario.

### Hypothesis testing with RIDE

Across 10 trials of propagation with *viral variant A* followed by hypothesis testing with RIDE given *Partial Reporting*, the difference between the number of expected minus observed passenger-to-driver rideshare infections was 16.7 (range -54.4 to 78.8) for *viral variant A* without masks or superspreaders; -61.0 (range -130.3 to 0.2) for *viral variant A with masks* without superspreaders; and 294.9 (range 224.8 to 371.4) for *viral variant B* without NPIs or superspreaders (all adjusted p-values ≤ 0.001). The results were qualitatively similar in the *Full Reporting* scenario, with greater differences, less variation, and a higher level of significance (Figure 3).

**Figure 3.**
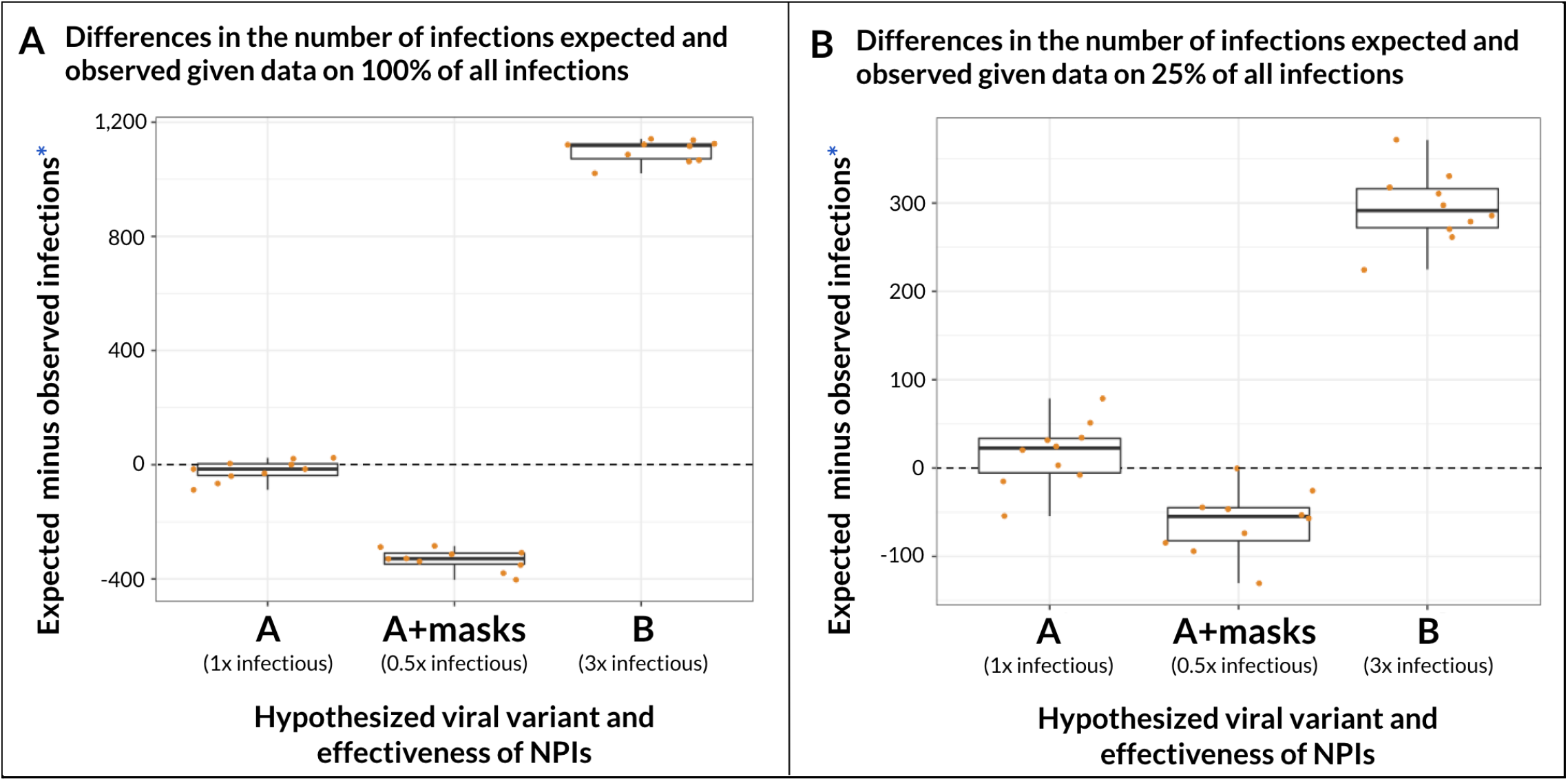
Differences given 10 trials in the number of passenger-to-driver infections expected and observed in the simulation based on analysis with different hypotheses of SARS-CoV-2 transmission, according to the percent of infections reported. Variability in results given analyses of 10 simulated synthetic datasets resulting from SARS-CoV-2 propagation in Los Angeles when true propagation conditions correspond to *viral variant A* with no masks and no superspreaders. For each simulated dataset, and with access to only diagnosed infections, the epidemiologist assesses the difference in the expected number of passenger-to-driver infections given analysis with hypothesized parameters (assuming either *viral variant A, viral variant A with masks*, or *viral variant B*) minus the adjusted number of observed rideshare infections in the network. Each dot within a box-plot represents analysis results with the given hypothesis for one round of Los Angeles simulation and analysis. Box-plot midline represents the median of analysis results across the 10 trials, box edges show interquartile range, and whisker tips show the minimum and maximum result values. *Observed infections adjusted to account for undiagnosed infections and baseline “false positive” transmissions (Supplementary IV).

For the secondary investigation comparing analytical results given simulated propagation of *viral variant A with superspreaders* instead of *viral variant A*, the mean observed number of drivers that infected exactly one passenger was 9.2 (range 29 to -6) lower than expected and the mean number of drivers that infected exactly two passengers was 1.8 (range -8 to 7) higher than expected for *Partial Reporting*. The results were qualitatively similar in the *Full Reporting* scenario, with greater differences (Figure 4). In both the *Full* and *Partial Reporting* scenarios, the combined data from the ten trials led to a significant difference in the distributions (*Full Reporting* p < 0.001, *Partial Reporting* p < 0.05).

**Figure 4.**
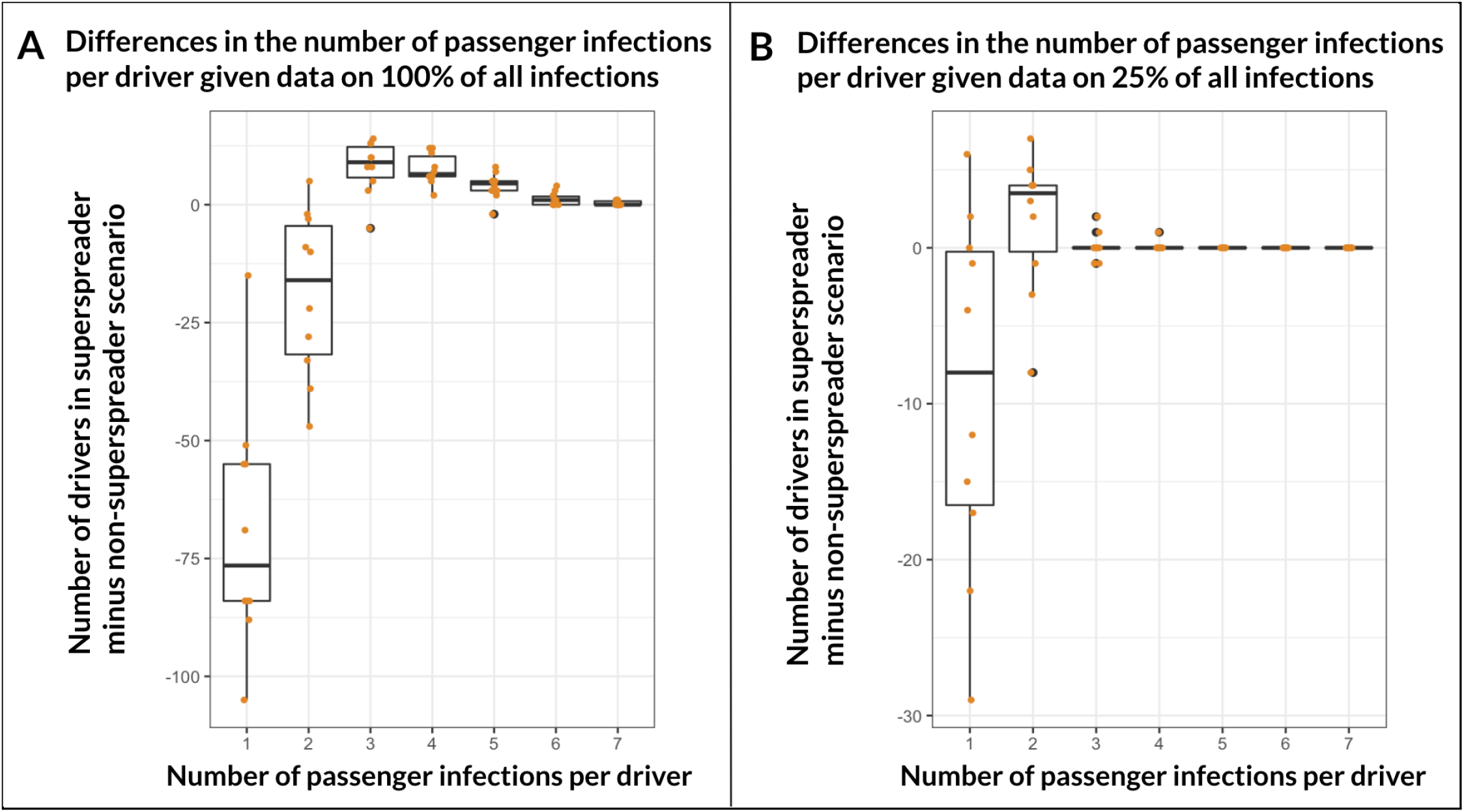
Differences in the number of passenger infections per driver in simulations with and without occasional superspreaders, according to the percent of infections reported. Variability in the differences between results given analyses of 10 simulated synthetic datasets resulting from SARS-CoV-2 propagation of *viral variant A* in Los Angeles given either a population with homogeneous infectivity (baseline, non-superspreader scenario) or asymmetric infectivity (superspreader scenario, where 5% of infected individuals are superspreaders). Box-plot midline represents the median of analysis results across the 10 trials; box edges show
interquartile range; whisker tips are minimum and maximum values; and black dot shows outliers, as
specified by Tukey.

## LIMITATIONS

The primary limitation of this work is the use of synthetic data. We do not study potential systematic infectivity differences between diagnosed and undiagnosed individuals, nor possible non-uniform diagnostic testing and reporting rates between drivers and passengers, nor between neighborhoods within cities. RIDE depends on estimates of the fraction of all cases that have been diagnosed and requires infected individuals’ symptom onset time relative to diagnosis, which may not be consistently recorded. However, this is a proof-of-concept whereas access to data from multiple cities along with detailed participant characteristics may be sufficient to overcome or mitigate these limitations.

## DISCUSSION

Using synthetic data generated based on empirical data from Los Angeles ridesharing and COVID-19 infections, we show that analyses of rideshare data may allow for the effective identification of the emergence of new strains of SARS-CoV-2 and the study of their transmission characteristics. Our analyses show that such an approach may be effective when as few as 25% of infectious individuals have been identified and suggest that further investigation is warranted for the development of such a system at the national scale and with access to more detailed data.

We demonstrate that research-based estimates for the range of SARS-CoV-2 transmission parameter values leave significant uncertainty in transmission modeling, leading to substantial variability in probability-of-infection predictions for a typical rideshare interaction. We demonstrate the ability of RIDE to differentiate between values within this estimated range, thereby enabling measured appraisal of: the emergence of more infectious viral strains; the effectiveness of Uber and Lyft’s nationwide mask mandates for all drivers and passengers; and the presence of superspreaders in the population. Straightforward expansions of RIDE could be used to evaluate the effectiveness of vaccines against newly emerging viral strains as well as to estimate their transmission characteristics such as patient infectivity relative to symptom onset; the effectiveness of other NPIs beyond simply masks; the prevalence and dynamics of passenger-to-passenger infections due to shared surface contact and leftover aerosols in the vehicle; and whether vaccinated individuals can still spread the virus. These models paired with the communication features of rideshare platforms could facilitate a largely-automated “radar” for contact tracing and targeted testing.^5^

Enhanced understanding of SARS-CoV-2 transmission dynamics through the use of mobile technology enjoys wide public and private support. Tens of millions of people have adopted technology developed by companies such as Apple and Google that use Bluetooth for a rapid notification system of potentially exposed users based on anonymously collected proximity data.^27–29^ However, these systems depend on probability-of-infection interaction modeling that is hindered by variable settings and uncertainty about SARS-CoV-2 transmission characteristics.^30^ Better estimates of SARS-CoV-2 parameters derived from interactions in more standardized settings could inform such efforts.

The US Center for Disease Control and Prevention may have grounds to require rideshare dataset access so that it can be merged with the list of positive COVID-19 cases, vaccination records, and other relevant data. The case has been made for digital disease surveillance that maintains considerations of ethics and patient privacy.^31^ Large rideshare companies such as Uber and Lyft are already sharing data with public health officials for contributing tracing, but no standardized analytics framework is available to aggregate and derive insights from these data.^16^ While logistics—the details of which are beyond the scope of this article—for the merging of rideshare and infection data are complex, it could be accomplished in an anonymous fashion given careful data processing.

## CONCLUSION

The rideshare network of exposure is unlike any other, with tens of millions of potentially infectious connections between individuals worldwide. Unlike more general cellphone-based contact data, rideshare contacts occur in relatively similar conditions, are sporadic and demographically diverse, and are almost always the only node connecting potential transmission between individuals. As demonstrated via simulations of COVID-19 propagation through Los Angeles’ rideshare network, viral strains with differing SARS-CoV-2 transmission parameters lead to detectably different patterns of infections, even in the presence of limited diagnostic information. Analyses of rideshare data combined with diagnoses records could facilitate an automated, local approach for the detection and investigation of emergent novel strains and the transmission characteristics of SARS-CoV-2 and future pandemics.

## Data Availability

All data was generated via simulation and is available upon request.

## ACKNOWLEDGEMENTS

We would like to thank James Safranek for his contributions to the development of the mathematical model of rideshare viral transmission. We also thank John Tamaresis and the Stanford Data Studio consulting group for their feedback on the statistical methods presented in this paper. Some of the computing for this project was performed on the Sherlock High-Performance Computing cluster. We would like to thank Stanford University and the Stanford Research Computing Center for providing computational resources and support that contributed to these research results.

