## Supplementary Appendix for "A model to analyze rideshare data to surveil novel strains of SARS-CoV-2"

**Table of contents:**

I. Overview

II. Model of SARS-CoV-2 transmission

III. Simulated propagation of SARS-CoV-2 through the Los Angeles rideshare network

IV. Analysis of rideshare infection patterns with RIDE

### I. Overview

A summary follows of the steps used to evaluate the potential of the Rideshare Infection Detection (RIDE) model to detect novel strains of SARS-CoV-2 and test hypotheses about their characteristics (used to generate Figure 3). Details are in Supplementary sections II to IV below.

1. Stochastically generate synthetic rideshare data for a city 1/200th the size of Los Angeles (LA) from March 17th to October 3rd (**Supplementary III**)
2. Randomly assign baseline infections to drivers and passengers based on historical LA infection rate estimations and trends. Record infection status, symptom onset time, and diagnosis status for each infected passenger and driver (**Supplementary III**)
3. Propagate rideshare transmissions using a mathematical model of rideshare transmission, and assuming that true transmission parameters correspond to *viral variant A* (details of mathematical model in **Supplementary II**)
    a. When an infection results (based on a draw from a Bernoulli distribution with the given probability of infection), assign symptom onset time and diagnosis status to the infected passenger or driver (**Supplementary III**)
4. Analyze the resulting infection pattern two times, assuming either *Full Reporting* or *Partial Reporting* conditions. Calculate potential transmission probabilities first assuming that infection parameters correspond to *viral variant A*. Specific steps include:
    a. Re-run the same code as in step 3 but with the following modifications:
        i. Only consider passenger-to-driver infections
        ii. For *Full Reporting*, consider the possibility of propagation for all potential passenger-to-driver infections, but for *Partial Reporting*, only consider diagnosed infections (*Partial Reporting* assumes that symptom onset data is available for only 25% of infections)
        iii. Add each potential passenger-to-driver infection probability to a *Potential* distribution that corresponds to current parameter assumptions
        iv. Instead of propagating infections as in step 3, check whether each potential infectee had symptom onset in the subsequent 3-10 day window. If so, count one *Observed* infection (note, this result is <u>independent</u> of the hypothesized parameters: you get same number of observed infections given analysis with all 3 hypothesized parameters) (**Supplementary IV**)
5. Repeat step 4 above twice more, assuming transmission parameters correspond to *viral variant A with masks*, and then assuming parameters correspond to *viral variant B*
6. Repeat steps 1 through 5 above 200 times total & aggregate the *Potential* probability-of-infection distributions (6 total: *Full* and *Partial Reporting*, for each of the

three hypothesized parameter assumptions) and *Observed* infection numbers (2 total: *Full /Partial Reporting*). Together, these results represent analysis of hypothetical aggregated rideshare and COVID-19 diagnosis data on LA from March 17th to October 3rd

7. Adjust the number of *Observed* infections to account for the percentage of cases diagnosed and for the average number of "false positives" given the overall density of infections in the network (**Supplementary IV**)
8. Calculate the expected number of infections based on the *Potential* probabilities under each set of hypothesized parameters, and for *Full* and *Partial Reporting* respectively
    a. The number of expected passenger-to-driver infections = the sum of all of the *Potential* passenger-to-driver infection probabilities (**Supplementary IV**)
9. Repeat steps 1 through 7 above 10 times to represent 10 trials. To generate Figure 3 ("Differences given 10 trials in the number of passenger-to-driver infections expected and observed in the simulation based on analysis with different hypotheses of SARS-CoV-2 transmission, according to the percent of infections reported"), plot the differences in the number of observed and expected passenger-to-driver infections based on analysis with different hypotheses of SARS-CoV-2 transmission (**Supplementary IV**)

The steps to generate Figure 4, "Differences in the number of passenger infections per driver in simulations with and without occasional superspreaders, according to the percent of infections reported," are similar but have a few key differences. First, two sets of synthetic rideshare infection patterns are generated via two distinct rounds of simulated propagation, the first with *viral variant A* (which assume homogenous infectivity across infected individuals), and the second with *viral variant A with superspreaders* (which assumes 1 in 20 individuals are *5x* more infectious). Second, the number of *Observed* passenger infections per infected driver (e.g. $N_1$ drivers appear to infect exactly 1 passenger, $N_2$ drivers infect exactly 2 passengers, etc.) are tabulated for both synthetic datasets. Next, the differences between these $N_i$ values are calculated between the two analyses. To generate the Figure 4, this process is repeated 10 times and the variability between simulations is plotted. See **Supplementary II-IV** for details.

### II. Model of SARS-CoV-2 transmission

The foundation of the present model comes from earlier work.[1,2] For the convenience of the reader, a description is reproduced here: "Since any of the numerous virus particles ingested by a susceptible can trigger an infection, we model the infection process as a Poisson process with the total accumulated viral load as the rate of infection."[1] Let the total ingested viral load for a given infector-infectee interaction equal $\lambda_i$. Thus, the probability that the potential infectee will get infected during the interaction is:

$$P(\lambda_i) = 1 - e^{-(\lambda_i)} \quad (1)$$

The ingested viral load $\lambda_i$ is dependent on various factors such as the infectivity of the infector, the duration of the interaction, and the rate of decay of virus aerosols particles. For each infection scenario—passenger to driver **($i = 1$)** and driver to passenger **($i = 2$)**—we use SARS-CoV-2 parameter estimates from recent literature to develop unique equations to determine $\lambda_i$ functions for each scenario that account for scenario-specific interaction dynamics.

**Passenger-to-driver transmission:**

Given $t_S$ hours since a patient's symptoms began (with negative values representing number of hours before illness onset), a patient's magnitude of infectivity can be modeled with a gamma distribution. A study of 77 well-defined infector-infectee pairs estimated this gamma function to have shape $a$ of 2.12, scale $b$ of 1.45, and a left-shift $c$ of 2.3 days (to represent that a patient becomes infectious 2.3 days before symptom onset).[3]

$$i(t_S) = \frac{(t_S/24 + c)^{(a-1)} \cdot e^{-(t_S/24 + c)/b}}{b^a \cdot gamma(a)} \quad (2)$$

There is significant uncertainty in the distribution, with the 95% confidence intervals for the left-shift and the peak infectivity time ranging from 0.8 to 3.0 days and –2.0 to 0.2 days relative to symptom onset, respectively.[3]

The cumulative amount of virus deposited in the vehicle, $d_1(t_R, t_S)$ at time $t_R$ hours into a ride with an infected passenger who is $t_S$ hours since symptom onset can be calculated by integrating the infectivity $i(t)$ of an individual, with respect to time, multiplied by a factor $m_\alpha$ to represent the unknown rate for the magnitude of virus particle expulsion.[2]

$$d_1(t_R, t_S) = \int_{t_S}^{t_S + t_R} m_\alpha \cdot i(\tau)\, d\tau \quad (3)$$

The total virus ingested by a susceptible driver up to time point $\boldsymbol{t_R}$ in a ride interaction is represented by the integral $\boldsymbol{\lambda_1(t_R,t_S)}$ which assumes that at each point in the interaction, the driver ingests an unknown rate percentage $\boldsymbol{m_\beta}$ of the total virus present in the vehicle.

$$\lambda_1(t_R,t_S) = \int_0^{t_R} m_\beta \cdot d(\tau')\, d\tau' \tag{4}$$

Thus, the total amount of virus ingested by the potential infectee during a $\boldsymbol{t_R}$ hour rideshare interaction is a double integral that varies quadratically with respect to time for short interactions (less than 1 hour).

Note that the rate of the infector's virus particle expulsion $\boldsymbol{m_\alpha}$ is dependent on the viral strain of SARS-CoV-2, a variable factor as demonstrated by studies showing the increased infectivity of certain strains.[5] Additionally, both $\boldsymbol{m_\alpha}$ and the rate of the infectee's virus ingestion $\boldsymbol{m_\beta}$ are affected by the use of facemasks, and are thus dependent upon non-pharmaceutical interventions (NPIs).

The product of the constants $\boldsymbol{m_\alpha}$ and $\boldsymbol{m_\beta}$ is a strain- and NPI-dependent product variable (denoted $\boldsymbol{m_1}$) representing the magnitude of viral particle transfer per hour. Thus, the amount of virus ingested by a driver given a $\boldsymbol{t_R}$ hour ride with an infected passenger $\boldsymbol{t_S}$ hours since symptom onset simplifies to:

$$\lambda_1(t_R,t_S) = m_1 \cdot \int_0^{t_R} \int_{t_S}^{(t_S+\tau')} i(\tau)\, d\tau\, d\tau' \tag{5}$$

There is an additional term to account for the virus ingested by the driver *after* the passenger exits the vehicle either due to leftover aerosols or surface particles in the vehicle. However, given that the driver and passenger do not touch the same surfaces and that infection probability from in-person contact is significantly higher than indirect contact, for our model we do not include this term.

**Driver-to-passenger transmission:**

When considering a passenger that rides in a vehicle with an infected driver, the key differentiating factor is that the viral load in the vehicle will likely already be saturated and therefore not change significantly over the course of the ride duration. This is based on the assumption that, given that a driver typically remains in their vehicle over the course of a workday, the rate of virus expulsion by the infected driver minus the rate of viral death and evacuation due to ventilation will stabilize. Therefore, this level of saturation $\boldsymbol{m_\gamma}$ is an unknown magnitude dependent on the equilibrium between viral deposition and subsequent evacuation and decay. When considering how much virus is ingested by the passenger, $\boldsymbol{m_2}$ is a combination of $\boldsymbol{m_\gamma}$ and the percentage ingestion rate $\boldsymbol{m_\beta}$. Thus, the total virus consumed $\boldsymbol{\lambda_2(t_R,t_S)}$ by a passenger $\boldsymbol{t_R}$ hours into a ride with a driver that is $\boldsymbol{t_S}$ hours since symptom onset simplifies to:

$$\lambda_2(t_R, t_S) = m_2 \cdot \int_{t_S}^{(t_S + t_R)} i(\tau)\, d\tau \qquad (6)$$

Unlike in the passenger-to-driver transmission scenario, note that cumulative viral ingestion increases linearly for short interactions (<1hr) due to the constant viral load present in the vehicle.

**Transmission Function Summary:**

As is standard in the literature, probability of infection is modeled as a Poisson process with rate $\lambda_i$ equal to the cumulative ingested virus by the potential infectee.[1] Given a $t_R$ hour ride with an infector that is $t_S$ hours since symptom onset, we develop two $\lambda_i$ infection scenarios functions for potential (1) passenger-to-driver and (2) driver-to-passenger transmission. Across the two scenario-specific functions included in the mathematical model, there are three previously estimated and two completely unknown transmission parameters.

| Model Component | Infector-Infectee Passenger(P)-Driver(D) Scenario Relevance | Parameter Symbol | Parameter Description | Estimates from Literature (mean, 95% CI) |
|---|---|---|---|---|
| **Relative magnitude variables define the NPI-dependent and strain-dependent rate of viral transfer between infector and infectee** | P→D | $m_1$ | rate of virus particle transfer from infectious passenger to susceptible driver — virus particles per hour | *Not Available* |
| | D→P | $m_2$ | rate of virus particle transfer from infectious driver to susceptible passenger — virus particles per hour | *Not Available* |
| **Infector's infectivity relative to symptom onset is modeled with a shifted gamma distribution.** | P→D and D→P | **a** | Shape of gamma distribution | 2.12 [*CI Not Available*] |
| | P→D and D→P | **b** | Scale of gamma distribution | 1.45 [*CI Not Available*] |
| | P→D and D→P | **c** | Shift of gamma distribution (# of days patient is infectious before symptom onset) — days | 2.3 [0.8–3.0] |

**Table S1:** Previously estimated (***a***, ***b***, and ***c***) and unknown ($m_1$ and $m_2$) transmission parameters included in mathematical model of probability-of-infection for a given potentially infectious rideshare interaction.

For driver-to-passenger or passenger-to-driver transmission, probability-of-infection depends on the duration of the ride $t_R$ and the number of hours $t_S$ since the infector's symptom onset relative to the timing of potential transmission during the rideshare trip.

| Input Name | Input Description | Infector-Infectee Passenger(P)-Driver(D) Scenario Relevance |
|---|---|---|
| $t_S$ | Time between infector's **symptom** onset and rideshare trip — hours | P→D and D→P |
| $t_R$ | Duration of potentially infectious **ride** (in-person transmissions) — hours | P→D and D→P |

**Table S2:** Independent variable inputs for mathematical modeling of potentially infectious interactions within the rideshare network.

Thus, the final equations for probability-of-infection $P(\lambda_i)$ given total virus ingested $\lambda_i$ by a susceptible individual, is as follows:

$$P(\lambda_i) = 1 - e^{-\lambda_i}$$

$$\lambda_1(t_R, t_S) = m_1 \cdot \int_0^{t_R} \int_{t_S}^{(t_S + \tau')} i(\tau)\, d\tau\, d\tau'$$

$\lambda_1$: *passenger to driver*

$\lambda_2$: *driver to passenger*

$$\lambda_2(t_R, t_S) = m_2 \cdot \int_{t_S}^{(t_S + t_R)} i(\tau)\, d\tau$$

**Figure S1:** Transmission model to determine probability-of-infection given $\lambda_i$ ingested virus particles during a potentially infectious rideshare interaction with duration $t_R$ and symptom onset time relative to ride start time $t_S$. Patient infectivity is modeled with a gamma distribution ***i(t)***, as detailed by Eq 2.

For transmission from an infected passenger to a susceptible driver, $\lambda_1$ increases roughly quadratically with ride duration due to the increasing viral load present in the vehicle over the course of the ride. In contrast, for driver-to-passenger transmission, $\lambda_2$ increases roughly linearly with time and is dependent on a stable viral load present in the vehicle of an infected driver.

The relative magnitude of infectivity for a patient $t_S$ hours after symptom onset was modeled as a gamma distribution $i(t_S)$, and both the infector's rate of viral expulsion and the infectee's percentage ingestion rate are merged into a variable $m_i$. The use of NPI's such as facemasks, increased ventilation, and sanitization can affect viral expulsion rate, ingestion rate, and decay rate, and therefore can be represented by varying the magnitude of $m_i$ in each of the three infection scenarios.

**Parameters representing SARS-CoV-2 viral variants, NPI implementation, and superspreaders:**

The following table contains details for the transmission parameters used to represent hypothetical SARS-CoV-2 variants and propagation conditions.

| Model Component | Infector-Infectee Passenger(P)-Driver(D) Scenario Relevance | Parameter Symbol | Parameter Description | Viral variant A | | Viral variant B (no masks) |
|---|---|---|---|---|---|---|
| | | | | *baseline, no masks* | *with masks* | |
| **Relative magnitude variables define the NPI-dependent and strain-dependent rate of viral transfer between infector and infectee** | P→D | $m_1$ | rate of virus particle transfer from infectious passenger to susceptible driver — virus particles per hour | **1.2** | **0.6** | **3.6** |
| | D→P | $m_2$ | rate of virus particle transfer from infectious driver to susceptible passenger — virus particles per hour | **0.4** | **0.2** | **1.2** |
| **Infector's infectivity relative to symptom onset is modeled with a shifted gamma distribution.** | P→D and D→P | **a** | Shape of gamma distribution — unitless | **2.12** | **2.12** | **2.12** |
| | P→D and D→P | **b** | Scale of gamma distribution — unitless | **1.45** | **1.45** | **0.9** |
| | P→D and D→P | **c** | Shift of gamma distribution (# of days patient is infectious before symptom onset) — days | **2.3** | **2.3** | **3** |

**Table S3:** Assumed value for SARS-CoV-2 transmission parameter determine interaction probability-of-infection modeling for two hypothesized test cases: Previously estimated parameters (a, b, and c) and unknown, NPI-dependent, magnitude-of-viral-transfer parameters ($m_1$ and $m_2$) are respectively varied to represent hypothetical SARS-CoV-2 variants and propagation scenarios.

*Viral variant A* includes values for transmission variables most directly based on previous literature (outlined in Supplementary I). However, as noted, for the three parameters of the gamma infectivity curve there were significant ranges for the 95th confidence intervals. Thus, for *viral variant B* we change the shift ***c*** and scale ***b*** of the gamma infectivity function to shift the number of days a patient is infectious before diagnosis from 2.3 to 3 days, and the peak infectiousness from -0.7 to -2 days (high level representation of this change in Table 1; detailed change to gamma distribution parameters in Table S3).

To explore the impact of NPIs such as facemasks, for the hypothetical scenario of *viral variant A with masks*, all values of $\boldsymbol{m_i}$ are decreased two-fold to represent a 50% reduction in total viral exchange between infector and infectee due to them both wearing masks. Additionally, based on a recent publication detailing the emergent D614G SARS-CoV-2 strain, we increase all $\boldsymbol{m_i}$ variables 300% for *viral variant B* to represent a higher viral titer for infectious individuals relative to the baseline *viral variant A*.[5]

Finally, to represent a scenario where the infector population has infectivity asymmetry (hypothesis "*viral variant A with superspreaders*"), we design a separate hypothetical propagation scenario based off of *viral variant A*, but where 1 in 20 infectors is considered a superspreader. For these superspreaders, for each potentially infectious interaction we increase the resulting probability of transmission by 500% (in the few cases in which this results in probabilities greater than 100%, the probability is set to 100%). To keep roughly the same overall number of transmissions in the network, we slightly decrease (21.053% reduction → 78.947% relative to "baseline" infectivity) each of the calculated probability-of-infections for the 19 out of 20 non-superspreaders in the population. Note that this definition of superspreader implies an increase in the viral load for superspreaders, but it does not incorporate any changes or assumptions about the quantity or timing of their interactions.

Values of $\boldsymbol{m_i}$ for baseline *viral variant A* were chosen to obtain reasonable numbers for the total new diagnosed infections in LA due to rideshares from the period of March 17th to October 3rd (200 days): specifically, the total number of new diagnosed infections due to propagation of each respective variant on average does not exceed 0.5% of the total 264,414 COVID-19 diagnoses during the period in LA, as estimated by five rounds of test propagation.

### III. Simulated propagation of SARS-CoV-2 through the Los Angeles rideshare network

**Approximation of the Los Angeles rideshare network from March 17th to October 3rd:**

Data available online were used to generate a realistic rideshare network that incorporates much of the stochasticity present in the rideshare patterns of real-world cities.

The distribution of the number of rideshares trips per day per driver and per passenger as well as the average duration of rideshare trips were derived from data published by the New York City (NYC) Taxi and Limousine Commission and adapted to simulate the rideshare network of Los Angeles (LA) county.[6] While this abstraction is imperfect, we chose this option because the NYC Taxi and Limousine Commission publishes the most comprehensive data on rideshare patterns, but the more gradual wave of COVID-19 positive diagnoses in LA is a better representation of other major US cities (as opposed to the sharp spike of early cases in NYC). To scale from the 8.6E6 population of NYC to the 10E6 population of LA, we multiplied relevant values by 1.163.

To generate a realistic estimate of the rideshare network for LA from March 17th to October 3rd (200 days), we started based on the following data from the NYC Taxi and Limousine Commission (Table S4).[6]

| ***Inputs to generate a hypothetical rideshare network*** *(estimates based on New York City):* |
|---|
| Number of rideshare drivers = **64,000 drivers** (→ 0.744% of NYC's total 8.6E6 population are drivers) |
| Average days/month a driver works = **20.5 days** (→ 68.8% chance a driver works on a given day) |
| Average rides given on a day a driver works = **13.63 rides/day** (→ 9.38 rides/day overall average for all days of month) |
| Average ride duration = **17 min** (distribution data unavailable, but assumed to have a lognormal distribution with log mean 2.65 min and log standard deviation 0.6053319) |
| Average interim period between rides = **unavailable** (assumed to have overall average of 12 min represented by a lognormal distribution with log mean 2.47 min and log standard deviation 0.8523) |
| Average total rides per day = **600,000 rides** |

**Table S4:** Relevant data from the New York City Taxi and Limousine Commission was gathered and adapted to model the Los Angeles rideshare network.

Data on the percentage of people who are rideshare users (about 40% of individuals living in cities), as well as the proportions of passengers that are daily, weekly, monthly, versus less-than-monthly users, were based on a 2019 PEW research poll examining rideshare habits in

U.S. cities.[7] The riding frequency of each of these passenger types (Table S5), was estimated using the PEW research poll and fine tuned in order to ensure that the total number of rides for a given day across all passengers' with their respective riding frequencies lined up with the total number of daily rides (~0.174 rides per rideshare user per day according to the NYC Taxi and Limousine Commission). These percentages and frequencies were assumed to be the same for LA (before adjusting for reduced quarantine utilization).

| | Percent of Rideshare Users | Rides / day | Rides/month |
|---|---|---|---|
| **daily riders** | 6.5% | 1.2 | 36 |
| **weekly riders** | 19% | 0.25 | 7.5 |
| **monthly riders** | 47% | 0.08 | 2.4 |
| **less-than-monthly riders** | 27.5% | 0.022 | 0.66 |

**Table S5:** Data from a 2019 PEW survey was incorporated to model the Los Angeles rideshare network.

We represent the rideshare network with a sparse three-dimensional tensor ("*rideshareNetwork*"), where each row represents a ride (with *Ride Number* corresponding to the number of that row) and each column represents a specific driver (with *Driver Number* corresponding to the number of that column). The start time of each ride is saved in layer one and the corresponding ride duration is saved in layer two of the *rideshareNetwork* tensor.

For each driver and for each day in the duration of the simulation, a draw from a Bernoulli with probability of success equal to 68.8% (see Table S4) determines whether the driver works that day. If they do, a start time for that day is determined by drawing from a normal distribution with mean 12:00 p.m. and standard deviation (SD) 2 hours. Next, the number of rides the driver gives that day is determined by drawing from a normal distribution (mean 13.63 rides and SD 3.5, rounded to nearest whole number). The code then builds the driver schedule into the *rideshareNetwork* tensor by looping through their rides that day and alternately determining ride durations (mean 17 minutes; distribution data not available, but estimated to have a lognormal distribution with log mean 2.65 and log SD 0.6053319) and interim periods between rides (average and distribution data not available, but assumed to have mean 12 minutes with a lognormal distribution with log mean 2.47 min and log SD 0.8523). Each row in the tensor under that driver column records the start time (tensor layer 1) and duration (tensor layer 2) for the given ride number. These steps are repeated 200 times for the first driver to represent each of the 200 days in the simulation, and then again fully repeated for each driver in LA. It is a sparse tensor because each row represents only one ride, so therefore the majority of the *rideshareNetwork* tensor remains empty.

For the 200 day window simulated, we incorporate the previously estimated 75% reduction in rideshare utilization due to the ongoing pandemic.[8] This 4-fold reduction is implemented with a 2-fold reduction to the probability of a driver working on a given day (reduced to 34.4%) and a

2-fold reduction to the average number of rides per day when a driver works (reduced to 6.815). This also corresponded to a 4-fold decrease in the average number of rides per day for each type of rideshare user (daily, weekly etc.).

To assign a *Passenger Number* to each *Ride Number* in the *rideshareNetwork* tensor, we generated a *passengerInRide* vector where the index positions of the vector correspond to a *Ride Number* (in the order they appear in the rideshare matrix) and each of the values in the vector correspond to a *Passenger Number*, representing the passenger in that ride. We incorporate the distribution of daily, weekly, monthly, and less-than-monthly riders into this vector, and finally we randomize the order of these *Passenger Numbers*.

To reduce computational complexity, the population of LA and the corresponding rideshare volumes are broken down into 200 smaller networks, each representing rides within one of 200 neighborhoods with a population of 50,000 people. Results from each of the 200 neighborhoods are aggregated later in the simulation analysis to represent data for all of LA. We assume that this parallelized analytical method does not impact overall results.

**Assigning SARS-CoV-2 infection and illness onset times to random passengers and drivers:**

For both passengers (i.e. riders) and drivers, hypothetical infection and diagnosis data is stored respectively in a *passengerPositiveMatrix* and *driverPositiveMatrix* with dimensions [rows ***x*** columns ***x*** depth] = [total number passengers/drivers ***x*** 2 ***x*** 2]. The methods by which these matrices are filled are detailed shortly, but this paragraph focuses on the type of infection data that is stored in the row, column, and depth dimensions. The index of each row corresponds to a *Passenger Number* or *Driver Number*, respectively, a unique identifier for each passenger and driver represented in the simulation. Column one stores infection and diagnosis status, with a 0 meaning the patient was not infected with COVID-19 during the simulated time period, a 1 meaning the patient was infected at some point during the simulation and diagnosed, and a 2 meaning the patient was infected during the simulation but not diagnosed. Column two stores the time of symptom onset if the patient was infected during the simulation. The first layer (depth = 1) of this matrix tracks baseline infections (infections that are initially randomly assigned to passengers and drivers), and the second layer (depth = 2) tracks "first degree" rideshare-acquired infections (simulated infections that passengers and drivers acquire from sharing rideshares with baseline infected individuals).

We estimate that, on average, the process of noticing symptom onset, scheduling a test, and getting tested typically takes about three days. With regards to simulated viral propagation, the amount of this shift is arbitrary, but means that the 264,414 positive diagnoses (diagnosis reporting corresponds to date of test sample collection) in LA from March 20th to October 6th correspond to about 264,414 instances of symptom onset during the window of time three days prior: Our simulation therefore represents LA during the 200 day window from March 17th to October 3rd. It is assumed that the COVID-19 diagnoses data collected by governmental agencies would consistently have information on symptom onset time relative to viral test date, as this is the data that is needed to calibrate individuals' gamma infectivity functions.

We approximate the time distribution for these 264,414 instances of symptom onset via a normal distribution (see actual and modeled data; Figure S2, Panels A and B).[9] Based on recent research suggesting significant infection due to transmission from undiagnosed individuals, it was assumed that the baseline infection rate during this time period represented only 50% of the actual infections in the population.[10,11] Therefore, the 2.64% baseline diagnosis rate in Los Angeles is assumed to correspond to a 5.29% baseline infection rate during this time period.

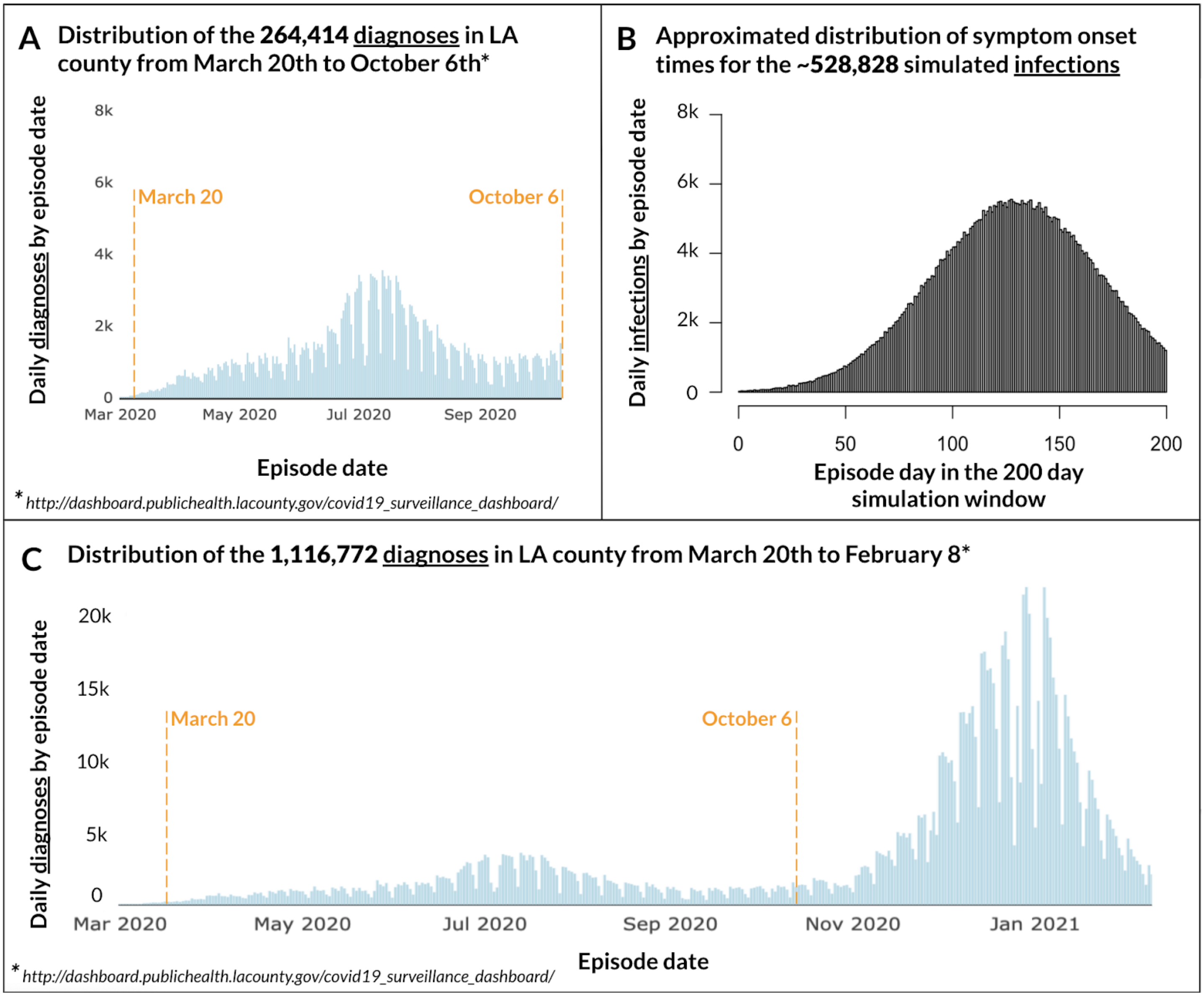


**Figure S2:** Comparison of empirical diagnoses data from Los Angeles County Public Health[9] between March 20th and October 6th (Panel A) and approximated symptom onset time data used for the 200 day window simulated (Panel B). Overall distribution of LA County diagnoses until February 8 shown for comparison, but not used for simulation (Panel C).

For each passenger and driver in the simulation, baseline infection status is determined via a draw from a Bernoulli distribution with a 5.29% probability of success. If positive, symptom onset time is assigned by drawing from a normal distribution with a mean of 130 days and an SD of 40 days (Figure S2, Panel B). If a calculated symptom onset time fell outside of the 200-day simulation window, it was recalculated randomly until a value between 0 and 200 was drawn.

Note that while LA has one of the greatest cumulative case counts in the world, for the chosen 200-day simulation window we deliberately excluded the more recent, and much larger, second wave of cases. Therefore, our simulation considered less than one quarter of the total cases in LA (Figure S2, Panel C). Although not including the second wave of cases greatly reduces the power of subsequent analyses, we made this decision such that our subsequent analyses and proof-of-concept would be applicable to other cities in the US. For comparison, as of March 18, 2021, there have been more than 10 US cities with at least 264,414 cumulative COVID-19 diagnoses (the number of diagnoses in LA for our selected 200-day simulation).[12]

**Representing *Full Reporting* and *Partial Reporting* scenarios:**

Throughout the analyses detailed in Supplementary IV, we consider two sets of hypothetical conditions: *Full Reporting* and *Partial Reporting*. For *Full Reporting*, the diagnosis status for every infection in column one of both the *passengerPositiveMatrix* and *driverPositiveMatrix* are marked as 1, meaning that 100% of infections are diagnosed and have corresponding symptom onset time data available. While this level of infection reporting was not the reality in LA during this time period, we consider this hypothetical scenario to represent the potential power of such analyses for future pandemics when governments may be better equipped to more rapidly scale testing capacity. For *Partial Reporting*, for each infection we determine diagnosis status via a draw from a Bernoulli distribution with probability of success equal to 25%. For the 75% of infections without available data in the hypothetical *Partial Reporting* condition, the missing data could be due to a combination of incomplete logs for symptom onset time data, lack of available testing for infected symptomatic individuals, or asymptomatic infected individuals who are not tested. Note that *Partial Reporting* means that 1 in 4 infections that are simulated during this time period in LA are diagnosed (we propagate from an assumed ~528,828 baseline infections, see above); However, *Partial Reporting* represents a scenario in which epidemiologists are able to gather data on about 50% of the 264,414 diagnoses reported in LA during this time period, which were assumed to be half of the true infections in the population.

For all simulated SARS-CoV-2 propagation described in this section (Supplementary III), we always propagate potential transmission from *all* infected individuals represented in the network, regardless of whether or not they were diagnosed. Diagnosis status is determined and recorded (via a draw from a Bernoulli distribution) for both assigned baseline and propagated rideshare-acquired infections because this information is necessary to subsequently analyze the resulting rideshare infection patterns when we assume *Partial Reporting* conditions (details in Supplementary IV).

**Simulated propagation to generate synthetic data later used to test RIDE's ability to detect differences in SARS-CoV-2 infectivity (relevant to Figure 3):**

We then propagate infections by applying the mathematical model of transmission (developed in Supplementary II) in order to generate synthetic data representing hypothetical SARS-CoV-2 transmission patterns within the LA rideshare network. For the first round of propagation (used to generate the synthetic data analyzed for Figure 3), we assume that the true viral variant and transmission conditions correspond to *viral variant A* (detailed in Table S3). Thus, for this propagation calibrate the relevant transmission functions (see Figure S1) with the SARS-CoV-2 transmission parameters for *viral variant A*.

Next, we determine all the possible *Ride Numbers* that contained a passenger who was infected at some point during the time window simulated. We create two additional vectors: *positiveRideNum* is used to record all *Ride Numbers* that may have contained an infectious passenger, and *positiveRideTimePatientInfected* is used to record symptom onset time for the infected passenger corresponding to the given *Ride Number*. To generate the values for these two vectors, we cycle through the *passengerPositiveMatrix* and consider every infected passenger (regardless of diagnosis status). For each positive case, we cycle through the *passengerInRide* vector and, for every match, we add the associated *Ride Number* to *positiveRideNum* and the associated symptom onset time to *positiveRideTimePatientInfected*.

We next consider all possible passenger-to-driver infections. For each potentially infected *Ride Number* in *positiveRideNum*, we check each column of the *rideshareNetwork* tensor to determine the start time of the ride. If the symptom onset time for the passenger associated with that ride minus the start time of the ride is greater than the gamma infectivity shift value *c* (equal to 2.3 days for *viral variant A*) or less than 8 days, it means that the ride was outside of the passenger's infectious window, and therefore that *Ride Number* is skipped and we proceed to the next value in *positiveRideNum*. However, if the ride start time was within the passenger's infectious window, then we use the ride duration time and passenger's relative symptom onset time to calculate a probability of the driver's infection according to equation 1 in Figure S1, and assuming true transmission parameters correspond to *viral variant A*. We determine infection via a Bernoulli trial with success equal to the corresponding infection probability. Given success, we determine diagnosis status with a separate Bernoulli draw with probability of success equal to the assumed diagnosis rate (100% or 25% for *Full* and *Partial Reporting*, respectively). We additionally determine a symptom onset time via a draw from a normal distribution with mean 5.2 days and SD 1.2 days.[3] Symptom onset time and infection/diagnosis status are recorded in layer 2 of the *driverPositiveMatrix*.

Finally, we consider all possible driver-to-passenger infections. For each infected driver, we first determine which of their rides occurred during the driver's infectious window (defined as in prior paragraph). For each of these rides when the driver was potentially infectious, we use the ride duration time and driver's relative symptom onset time to calculate a probability for the passenger's infection according to equation 2 in Figure S1, and assuming true transmission parameters correspond to *viral variant A*. Following the methods described in the prior paragraph, for any resulting infections we record the driver's diagnosis status and the driver's symptom onset time in layer 2 of the *passengerPositiveMatrix*.

For both passenger-to-driver and driver-to-passenger infections, we also tabulate the cumulative number of diagnosed rideshare infections.

**Simulated propagation to generate synthetic data later used to test RIDE's ability to detect the presence of superspreaders (relevant to Figure 4):**

To generate the synthetic data analyzed for Figure 4, two sets of synthetic rideshare infection patterns are generated via two distinct rounds of simulated propagation: first with *viral variant A* and second with *viral variant A with superspreaders*. As defined in Supplementary III, while *viral variant A* transmission parameters assume homogenous infectivity across all infected individuals, *viral variant A with superspreaders* transmission parameters assume infectivity asymmetry where 1 in 20 individuals are considered superspreaders and are 500% as infectious, and 19 out of 20 are 78.947% as infectious as compared to *viral variant A* (this reduction in infectivity for non-superspreaders keeps the overall number of rideshare infections about the same).

These two rounds of propagation follow much of the same steps described above, with a few modifications. First, the *passengerPositiveMatrix* and *driverPositiveMatrix* have two additional columns beyond the original columns 1 and 2; column 3 records the magnitude of overall relative infectivity for that individual (thereby representing whether they are a non-superspreader or superspreader); and column 4, which is not used until the analysis phase (Supplementary IV), is used to tally the number of individuals that the individual appears to infect during the simulation. Second, while all infected individuals with *viral variant A* have a 1 in column 3, for *viral variant A with superspreaders* when baseline infections are assigned to passengers and drivers, a draw from a Bernoulli distribution with a probability of success of 5% determines whether the individual is a superspreader, and their corresponding magnitude of infectivity is recorded in column 3. The steps of simulated propagation proceed as in the prior section, except after calculating each probability of potential rideshare infection and before drawing from the Bernoulli distribution to determine transmission, the infection probability is multiplied by column 3, the potential infectee's magnitude of infectivity.

### IV. Analysis of rideshare infection patterns with RIDE

Having generated hypothetical synthetic data representing SARS-CoV-2 rideshare transmission patterns in LA, we then analyze the data as if we were an epidemiologist unaware of which infections in the network were baseline versus rideshare-acquired, and also without access to data on undiagnosed infections. For both of the analyses used to produce Figure 3 and Figure 4, we repeat all stages two times to represent the analytical results the epidemiologist might discern given hypothetical conditions of *Full Reporting* (data available on 100% of infections) versus *Partial Reporting* (data available on 25% of infections). For the purposes of demonstrating the feasibility of Rideshare Infection Detection (RIDE), we specifically determine the ability of RIDE to differentiate between a number of predefined, hypothesized parameter sets representing specific viral strains and transmission scenarios.

**Baseline analysis to detect differences in SARS-CoV-2 infectivity (relevant to Figure 3):**

For the first set of synthetic data, which was generated with simulated propagation of *viral variant A*, we determine the ability of an epidemiologist using RIDE to retrospectively differentiate the correct set of transmission parameters given three potential hypotheses with different levels of infectivity: *viral variant A*, *viral variant A with masks*, or *viral variant B* (see Supplementary II, Table S3 for details). Overall, this analysis with RIDE includes the following steps repeated two times, once for *Full Reporting* and once for *Partial Reporting*: 1) We count observed infections in the network, but have to adjust this number to account for *Partial Reporting* and "false-positives" (details below); 2) We then use the same set of analytical steps for each of the three hypotheses to generate an expected number of rideshare-acquired infections given each set of assumed transmission parameters and considering all potentially infectious rideshare interactions in the network; 3) For each hypothesis, we take the respective differences between the projected number of expected infections for that hypothesis and the number of observed infections; this difference is expected to be closest to zero when calculated given the correct hypothesis; 4) We repeat this whole process 10 times—representing the impact of natural variability in LA's rideshare network and the stochasticity of SARS-CoV-2 transmission—to examine the consistency of RIDE's ability to identify the correct hypothesis.

For this stage of analysis, we choose to focus on exclusively analyzing possible passenger-to-driver infections. As detailed in Supplementary II, even for two identical scenarios (same SARS-CoV-2 variants, NPI use, ride duration, and timing of the potential infector's symptom onset relative to the rideshare trip), the probability of infection for a passenger-to-driver versus driver-to-passenger transmission is not equal. This is primarily because when a driver is infectious, virus particles build up in the car before the passenger enters the vehicle, thereby increasing relative driver-to-passenger transmission probability. The magnitude of this difference in the transmission parameters for these scenarios is currently unknown. Therefore, while the decision to focus on either passenger-to-driver versus driver-to-passenger infections is somewhat arbitrary, aggregated analysis of passenger-to-driver and driver-to-passenger infections is not currently possible because it introduces an additional unknown. Our decision to test the ability of RIDE to differentiate differences in infectivity while only considering passenger-to-driver infections is a conservative choice because, as incorporated in our simulated viral propagation, driver-to-passenger infections are relatively more prevalent. In future real-world use of RIDE, comparing the relative prevalence of these two types of infections given data from even just a handful of cities might allow determination of the relative magnitude of these transmission parameters, a relative mathematical relationship that we expect to remain somewhat constant across locations and time. Once this relationship is determined, then future analysis could likely aggregate analysis of both potential passenger-to-driver and driver-to-passenger infections, which would even further increase the power of RIDE.

For each of the three sets of hypothesized transmission parameters (each of which is subsequently referred to as the ***current hypothesis***), we start by calibrating the functions defining the mathematical model of transmission with the parameters corresponding to the *current hypothesis*.

Next, we use steps similar to those used for the original simulation of passenger-to-driver viral propagation, with a few key differences to account for the data to which an epidemiologist would have access to in the real world. When developing *positiveRideNum* (the list of all *Ride Numbers* that could be potentially infected by a passenger) and *positiveRideTimePatientInfected* (the time of the passenger's symptom onset corresponding to each of these rides), we consider only diagnosed infections. We also include diagnosed rideshare-acquired infections in addition to just the baseline infections that were used to simulate the propagation patterns (i.e. we check whether there was a 1 in column 1 of either layer 1 <u>or</u> layer 2 of *passengerPositiveMatrix*), even though in our simplified simulated propagation we did not include the possibility of passenger-to-driver transmission for passengers who had rideshare-acquired infections from drivers.

For determining which drivers might be infected in these potentially infectious rides, we again deviate from the original propagation steps once a probability of infection for a given potentially infectious interaction has been calculated. Rather than actually infecting the driver in the rideshare, we instead add the probability to a *Potential* distribution corresponding to the *current hypothesis*. Next, we check whether the potentially infected driver was diagnosed (either baseline or rideshare-acquired, corresponding to a 1 in column 1 of layer 1 <u>or</u> 2 of *driverPositiveMatrix*) and had symptom onset in the 1.5 to 10 day window following the interaction. If so, we tabulate this as an *Observed* passenger-to-driver infection. Note that the 1.5 to 10 day window captures 99.89% of the propagated **diagnosed** rideshare-acquired infections (which had symptom onset time assigned via a draw from a normal distribution with mean 5.2 days and SD 1.2 days). This is likely an overestimate in the real world due to COVID-19 testing appointment delays.

After cycling through all rideshare trips with a potentially infectious passenger, we have a *Potential* distribution of all possible passenger-to-driver infection probabilities, as well as a total *Observed* number of passenger-to-driver infections. We then repeat these steps for the other two hypothesized sets of transmission parameters to develop two additional *Potential* distributions of probabilities. Note that the *Observed* number of infections is the same under all three hypotheses. Finally, we repeat for both *Full Reporting* and *Partial Reporting*, resulting in a final of two distinct *Observed* numbers of passenger-to-driver infections and 6 distinct *Potential* distributions of passenger-to-driver infection probabilities.

Also remember that, as described in Supplementary III, propagation and analysis is all performed at the level of a city 1/200th the size of LA, and then analytical results (the *Potential* distributions and *Observed* number of infections) are aggregated over 200 trials to represent data on one city with the characteristics of LA.

**Adjusting analysis results to account for *Partial Reporting* (relevant to Figure 3):**

Next, for the results collected under the *Partial Reporting* condition, we have to make an adjustment to account for the differential impact of undiagnosed infections on detection of *Observed* and *Potential* transmissions. In summary, while *Partial Reporting* leads to a decrease in both the number of detected *Observed* and *Potential* transmissions, this decrease disproportionately impacts detection of *Observed* infections in the network. To demonstrate this in the simplest way possible, instead of considering *Partial Reporting*, we compare a scenario with symptom onset data for 100% of cases to a scenario with data on 50% of cases (Figure S3).

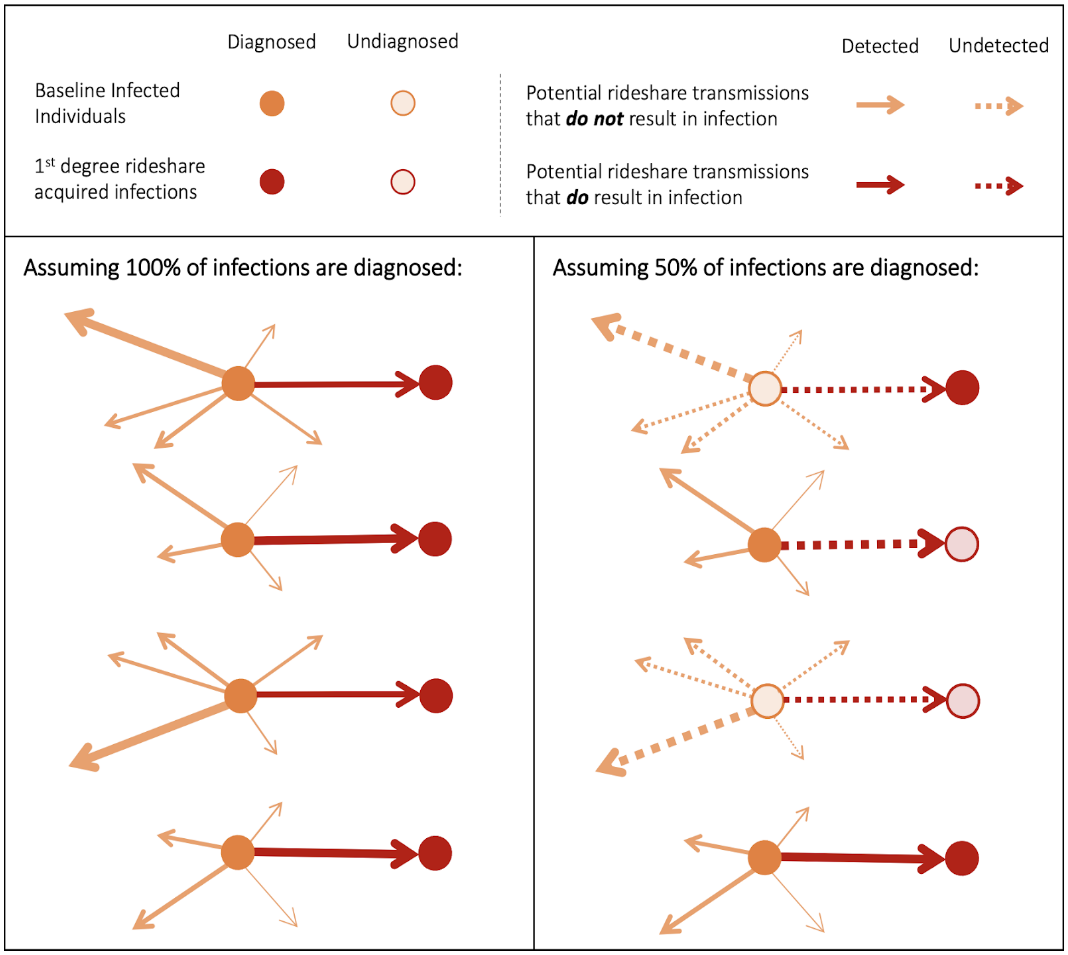


**Figure S3:** Simplified schematic of rideshare SARS-CoV-2 infection pattern analyses under two levels of reporting (when 100% or 50% of infections are diagnosed) to demonstrate the differential impact of undiagnosed infections on *Potential* transmission probabilities and *Observed* infections. Specifically, a ***p***% reporting rate for infections reduces the number of observable *Potential* infection probabilities by a factor of (1-***p***) and reduces the number of *Observable* infections by a factor of (1-***p***)².

As demonstrated in Figure S3, as data availability decreases, the number of detectable *Observed* infections decreases more rapidly than the number of detectable *Potential* rideshare transmissions. This is because, in order to identify an apparent rideshare transmission, both the infector and infectee need to be diagnosed, so the impact of the missing diagnosis data is compounded. In contrast, although there is a decrease in the total number of detectable *Potential* transmission as well, the impact of partial data is not compounded in the same way. Specifically, a reporting rate with symptom onset data for $\boldsymbol{p}$% of infections reduces the number of observable *Potential* infection probabilities by a factor of $(1\text{-}\boldsymbol{p})$ and reduces the number of *Observable* infections by a factor of $(1\text{-}\boldsymbol{p})^2$. To adjust for this differential impact via the simplest possible mechanism, we increase the number of *Observed* infections via multiplication by $1/(1\text{-}\boldsymbol{p})$.

**Raw difference in *Expected* minus *Observed* infections (relevant to Figure 3):**

After this adjustment to the *Observed* number of infections, we can then proceed with analysis. As previously described, the goal of this analysis is to correctly identify which of the three potential transmission hypotheses corresponds to the true set of parameters used to simulate viral propagation in the network. The next step is to convert each of the distributions of *Potential* transmission probabilities into an *Expected* number of infections. For both *Full* and *Partial Reporting*, and for each of the three hypotheses, we sum the probabilities of infection across all *Potential* rideshare interactions that could have resulted in infection. This sum of the expected values of the Bernoulli distributions represents the total number of *Expected* rideshare infections that would be expected to occur under each hypothesis given the quantity, duration and relative timing of all potentially infectious interactions within the rideshare network. For the correct hypothesis of *viral variant A*, the number of *Expected* infections should be about equal to the number of *Observed* infections in the network, and therefore the difference of *Expected* minus *Observed* should be close to zero; In contrast, for *viral variant A with masks* and *viral variant B*, because the hypothesized transmission parameters represent a less or more infectious variant/scenario as compared to the true scenario of viral propagation, then this *Expected* minus *Observed* difference would be expected to be negative or positive, respectively.

Due to the finite number of potential transmissions and the stochastic nature of SARS-CoV-2 transmission (given a draw from a Bernoulli), there is inherent variability in the number of rideshare infections. To capture this variability in addition to that which might occur due to fluctuation of the rideshare network (detailed in Supplementary III), we repeat the aforementioned series of propagation and analyses a total of 10 times. For each level of reporting (*Full*/*Partial)* and for each hypothesis, we then plot the 10 differences across the 10 trials of LA in order to examine the consistency of RIDE's ability to identify the correct hypothesis (Figure S4).

However, we found that under the correct hypothesis, these raw differences in the number of *Expected* minus *Observed* infections were consistently negative for both *Full* and *Partial Reporting*. After some consideration, we realized this was likely due to false-positives in the *Observed* distribution.

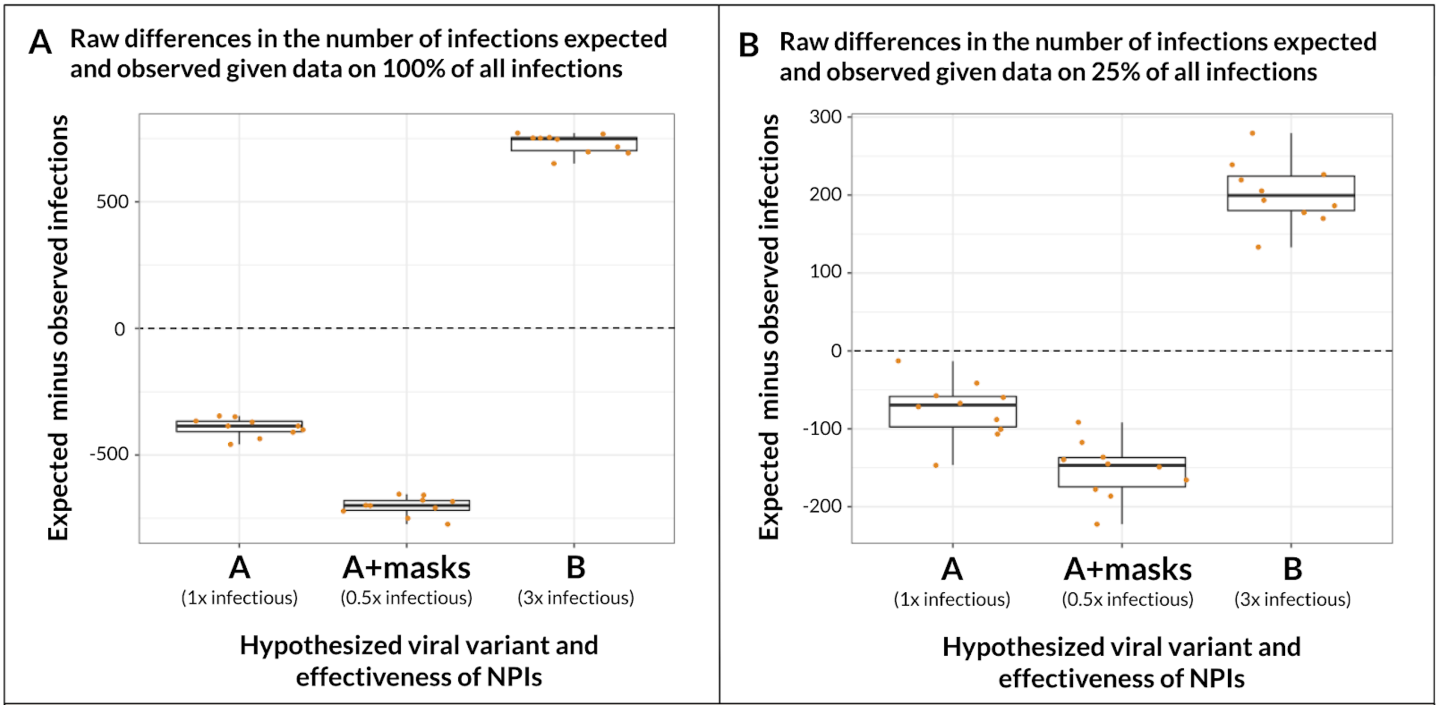


**Figure S4: Raw differences given 10 trials in the number of passenger-to-driver infections expected and observed in the simulation based on analysis with different hypotheses of SARS-CoV-2 transmission, according to the percent of infections reported.** Variability in results given analyses of 10 simulated synthetic datasets resulting from SARS-CoV-2 propagation in Los Angeles when true propagation conditions correspond to viral variant A with no masks and no superspreaders. Each dot within a box-plot represents analysis results with the given hypothesis for one round of Los Angeles simulation and analysis. Box-plot midline represents the median of analysis results across the 10 trials, box edges show interquartile range, and whisker tips show the minimum and maximum result values.

**Estimating and adjusting the number of "false-positive" *Observed* infections given rideshare network infection density (final step to generate to Figure 3):**

Because analysis of rideshare infection patterns must be performed without knowledge of which diagnosed infections are rideshare-acquired versus due to other causes (ie. "baseline infections," which are randomly assigned to initialize the simulated propagation), there is misclassification in some instances. Specifically, we incorrectly count a "false-positive" *Observed* infection when a potentially infectious individual takes a rideshare trip and does not transmit the virus to the other person in the vehicle, but the other person gets sick due to other causes and is diagnosed in the 10-day window following the rideshare trip. These "false-positive" transmissions can appear to occur from one baseline individual to another, from a rideshare-acquired infected individual to a baseline individual, from a baseline individual to a rideshare-acquired infected individual who actually got sick due to a different rideshare, or from a rideshare-acquired infected individual to another rideshare-acquired infected individual who actually got sick due to a different rideshare. The first and third of these cases are demonstrated in Figure S4.

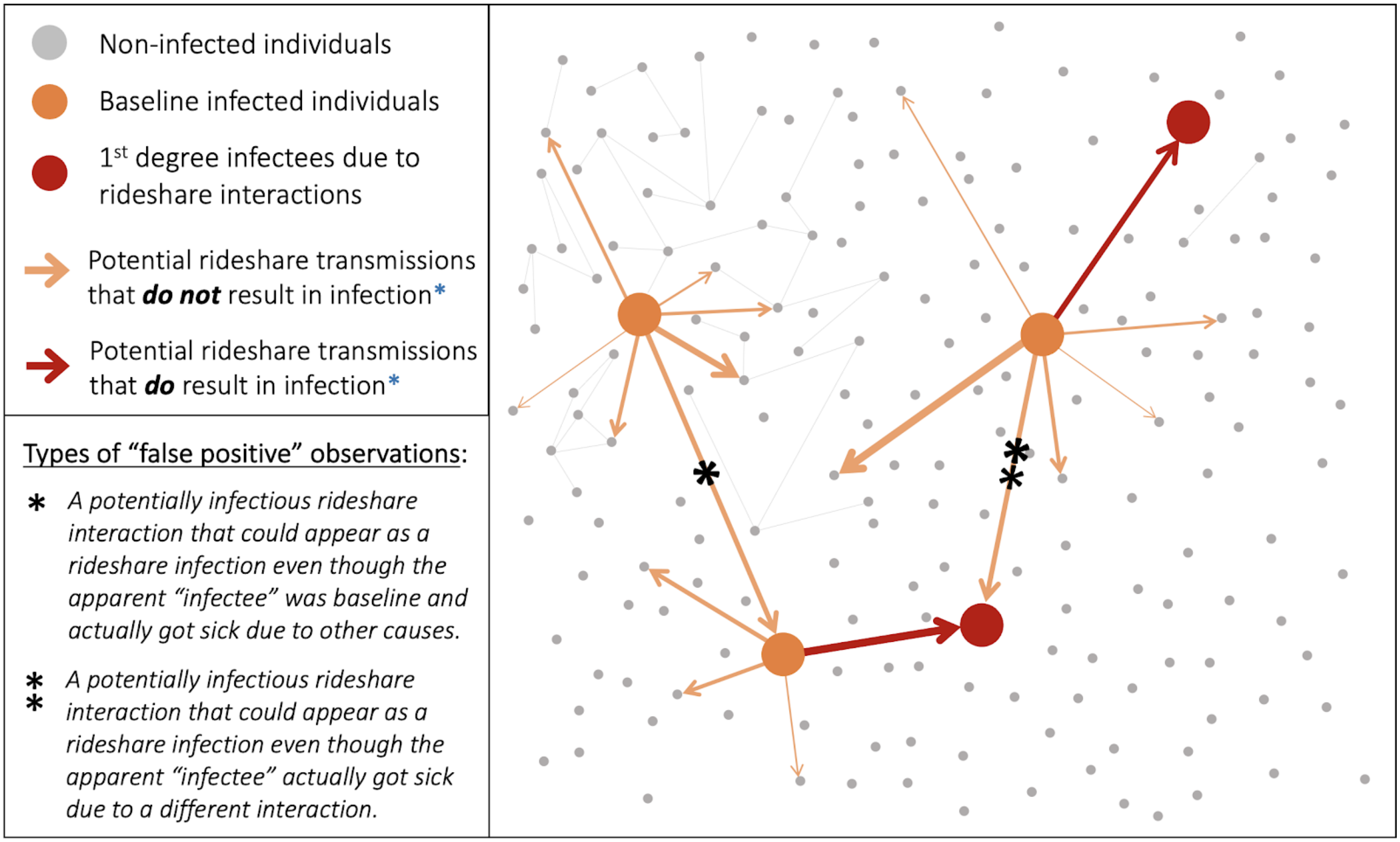


**Figure S4: Mock infection patterns within a rideshare network demonstrate the possibility of "false-positive" transmission observations**. An epidemiologist who cannot differentiate baseline (orange) from rideshare-acquired (red) infected individuals will falsely count *Observed* transmission connections between diagnosed individuals (specifically "*" and "**" in the figure) as the source of infection when in reality the potential "infectee" (at the end of each arrow) got sick from another cause.

These "false-positive" connections incorrectly inflate the number of *Observed* infections during analysis, thus leading to the negative differences measured for the correct *viral variant A* hypothesis in Figure S3. It is unfortunately impossible to classify which of the *Observed* infections are due to "false-positives;" however, it is still feasible to estimate their prevalence given the overall infection density in the network, and thus correct for the false inflation for the number of *Observed* infections.

To do this, we measure the expected prevalence of these "false-positives" given the overall infection density in the network via an additional round of simulations. In these simplified and altogether separate simulations, we reuse the same code as described above, but with three modifications:

First, instead of initiating the simulation with a baseline infection rate in LA of 5.29%, we also account for the additional numbers of drivers and passengers that are infected due to rideshares. Specifically, the 1,666.4 ride-share acquired infections on average in the simulated propagation correspond to an average of 1,047 additional passenger infections and 619.4 driver infections. Using these two values and estimations of the total numbers of rideshare passengers and drivers in LA, we calculate the additional probability of baseline-infection for each of these two groups. For this separate, simplified simulation to quantify "false-positive" prevalence, we add each of these respective additional percentages to the baseline infection probability when we assign baseline infections to passengers and drivers. Second, we altogether eliminate the simulated propagation phase. This results in a fake "simulated rideshare infection pattern" in which all infections are actually randomly assigned (ie. baseline-acquired). Third, when we proceed to the analysis phase, we do not measure *Potential* probabilities, but instead just count the number of *Observed* passenger-to-driver infections (with the same method as described above).

We repeated this analysis 4 times with both *Full* and *Partial Reporting* and counted an average of 370 and 92 "false-positive" passenger-to-driver connections (Figure S5).

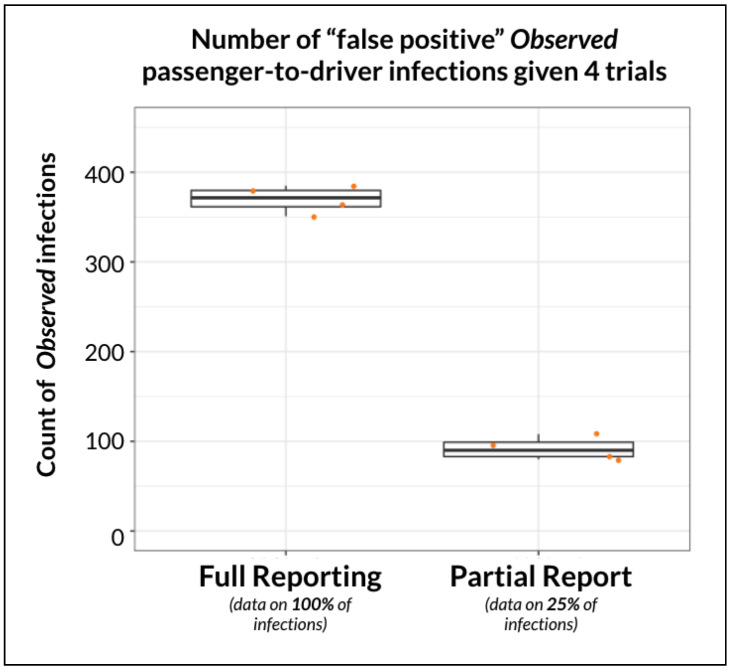


**Figure S4: Count of "false-positive" *Observed* passenger-to-driver transmissions across analysis of four trials given simulation with no viral transmission, and for *Full* and *Partial Reporting*.**

Note that, while the process of estimating the number of "false-positives" would be slightly different than what is described above for real-world data on diagnoses and the rideshare network, we believe that it would indeed be feasible. The calculations for the first of the simplified simulation modifications described above (increasing baseline infection probability for drivers and passengers) could be substituted by simply calculating the baseline infection rates for drivers and passengers in the aggregated rideshare and COVID-19 diagnosis data. Then, the epidemiologist could use the historic rideshare data combined with an algorithm similar to the code described here, randomly assign passenger and driver infections within the real rideshare network (ignoring the true diagnoses patterns), and then count the number of *Observed* "false-positive" infections. To achieve sufficient accuracy for these "false-positive" estimates, it may be necessary to account for differences in rideshare-use prevalence and disease prevalence among different sub-neighborhoods within each city. However, with access to both COVID-19 diagnosis records and rideshare network data, it would be possible to account for both of these potential clustering effects via localized "false-positive" estimations with neighborhood-specific sub-implementation of the random assignment strategy described above.

For the final step of analysis required to generate Figure 3, we account for these "false-positives" and adjust the raw results shown in Figure S3 by decreasing the number of *Observed* rideshare infections by 370 and 92 for *Full* and *Partial Reporting*, respectively. After this adjustment, we correctly observe that analysis with hypothesis *viral variant A* yields *Expected* minus *Observed* differences that are more consistently close to zero (see main manuscript for statistical analysis and Figure 3).

**Analysis to test RIDE's ability to detect the presence of superspreaders (relevant to Figure 4):**

In a separate, secondary investigation, we examine the potential ability of RIDE to measure the prevalence of SARS-CoV-2 superspreaders. To accomplish this, we analyze and compare results for the two synthetic rideshare infection patterns generated via propagation of either *viral variant A* or of *viral variant A with superspreaders* (see end of Supplementary III for simulation details).

Recall that we represent population infectivity asymmetry (hypothesis "*viral variant A with superspreaders*") via inclusion of superspreaders 1 in 20 infectors who is considered a superspreader, where we increase the probability of each potentially infectious interaction by 500%. Given this, we expect scenario *viral variant A with superspreaders* to have a higher number of individuals who infect 2+ individuals. To increase the likelihood of observing this effect, we focus on driver-to-passenger infections for this analysis because, given that drivers generally spend more time in rideshare vehicles compared to passengers, the odds of multiple rideshare transmissions during their infectious window is much higher.

For this analysis, we use a simplified strategy based on the methods described above to identify *Observed* infections in the network. For each *Observed* driver-to-passenger infection, instead of just adding it to a total count, we increase the count in column 4 of *driverPositiveMatrix*. Thus, we tabulate the respective number of driver-to-passenger infections associated with each infected driver. We perform this analysis for both synthetic datasets 10 times to represent 10 hypothetical LA counties, and repeat for *Full* and *Partial Reporting*.

Results on the distribution of passenger infections per infectious driver for three randomly selected trials are shown in Figure S5. Across these three trials for *Full Reporting*, there is consistently a greater degree of right skewness for the infection patterns generated with *viral variant A with superspreaders*. However, for *Partial Reporting* this difference is not always present (see Trial 3). Note that the results in Figure S5 include "false-positive" *Observed* driver-to-passenger infections.

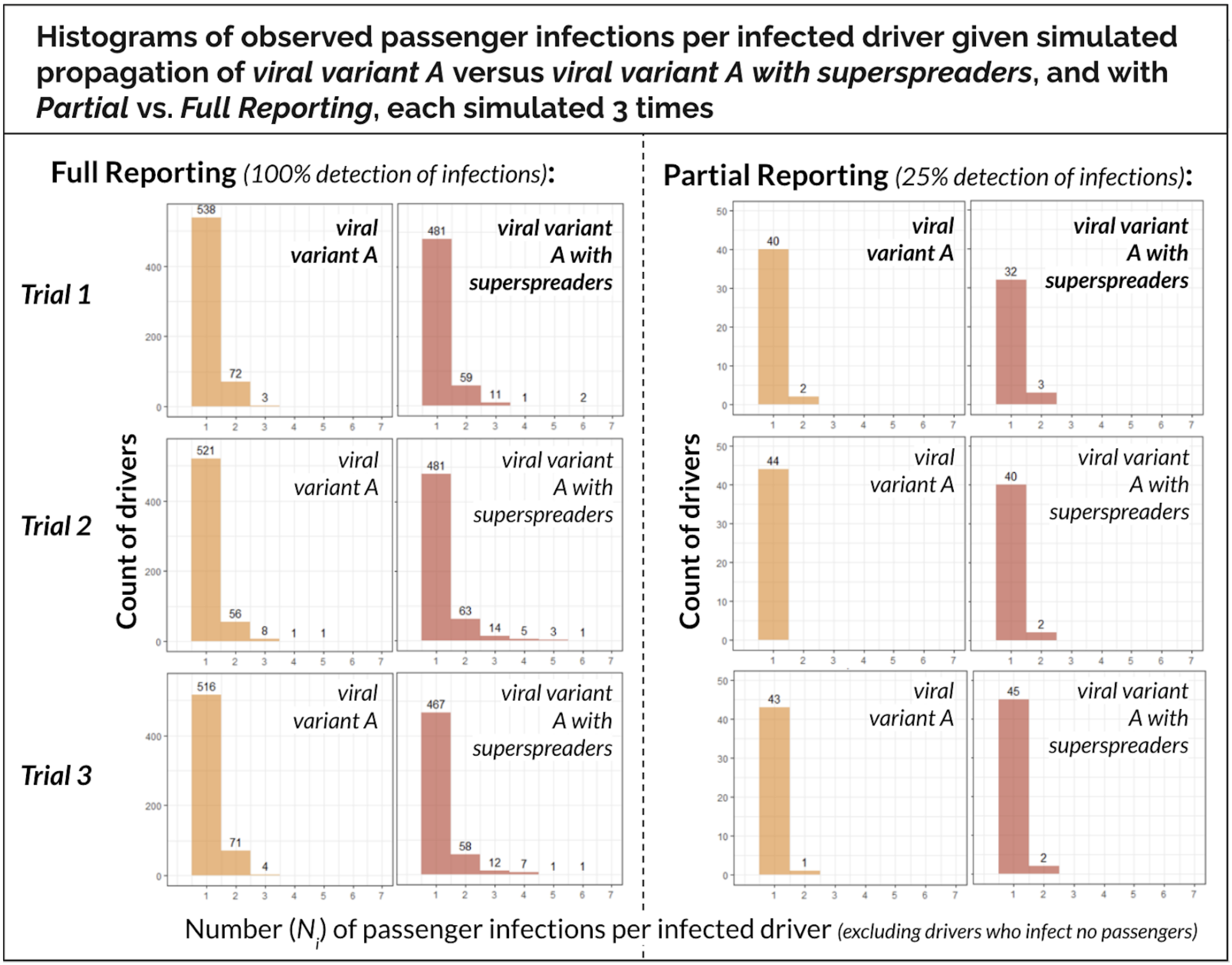


**Figure S5: Histogram results of passenger infections per infectious driver across three trials of simulated propagation of either *viral variant A* or *viral variant A with superspreaders*.**

As the final step to produce Figure 4, for each of the 10 trials we take the difference between the number of passengers infected per infectious driver observed in the *viral variant A* propagation scenario minus those observed in the *viral variant A with superspreaders* scenario (see main manuscript for statistical analysis and Figure 4). Note that this difference eliminates the problem of “false-positives” for this analysis because the pattern in the number of “false-positive” passenger infections per driver is expected to be on average equivalent for both scenarios.
